# The Health for Life in Singapore (HELIOS) Study: delivering Precision Medicine research for Asian populations

**DOI:** 10.1101/2024.05.14.24307259

**Authors:** Xiaoyan Wang, Theresia Mina, Nilanjana Sadhu, Pritesh R Jain, Hong Kiat Ng, Dorrain Yanwen Low, Darwin Tay, Terry Yoke Yin Tong, Wee-Lin Choo, Swat Kim Kerk, Guo Liang Low, The HELIOS Study team, Benjamin Chih Chiang Lam, Rinkoo Dalan, Gervais Wanseicheong, Yik Weng Yew, Ee-J Leow, Soren Brage, Gregory A Michelotti, Kari E Wong, Patricia A Sheridan, Pin Yan Low, Zhen Xuan Yeo, Nicolas Bertin, Claire Bellis, Maxime Hebrard, Pierre-Alexis Goy, Kostas Tsilidis, Harinakshi Sanikini, Xue Li Guan, Tock Han Lim, Lionel Lee, James D Best, Patrick Tan, Paul Elliott, Eng Sing Lee, Jimmy Lee, Joanne Ngeow, Elio Riboli, Max Lam, Marie Loh, John C Chambers

**Author notes:** Corresponding author: John Chambers, Lee Kong Chian School of Medicine, Nanyang Technological University, Level 18 Clinical Sciences Building, 11 Mandalay Road, 308232, Singapore. Contributed equally as the first authors.

## Abstract

Asian people are under-represented in population-based, clinical, and genomic research.^1,2^ To address this gap, we have initiated the HELIOS longitudinal cohort study, comprising comprehensive behavioural, phenotypic, and genomic measurements from 10,004 Asian men and women of Chinese, Indian or Malay background. Phenotyping has been carried out using validated approaches, that are internationally interoperable. Health record linkage enriches both baseline phenotyping and evaluation of prospective outcomes. The integrated multi-omics data include whole-genome and RNA sequencing, quantification of DNA methylation, and metabolomic profiling. Our data reveal extensive lifestyle, physiological, genomic, and molecular diversity between the distinct Asian ethnic groups, and the biological interconnectivity between functional layers. This includes characterisation of divergent patterns of genome regulation between Asian individuals, that correlate with differences in educational attainment, dietary quality, and adiposity, and which overlap transcription factors and DNA methylation sites linked to the development of diabetes and other chronic diseases. Our unique HELIOS Asian Precision Medicine cohort study represents a state-of-the art platform to enable biomedical researchers to understand the aetiology and pathogenesis of diverse disease outcomes in Asia, and to generate insights that have the potential to improve health outcomes for Asian populations globally.

## Introduction

Age-related chronic diseases such as diabetes, cardiovascular and respiratory diseases, cancer, and cognitive decline are leading causes of morbidity and mortality across all regions of the world^3^. Driven by demographic transitions, increasing urbanisation, and adoption of unfavourable lifestyle choices, the Asia Pacific region in particular is facing a rapid increase in chronic disease burden that contrasts stable or falling disease rates in Europe and North America^4^. There are already 296 million Asian people living with diabetes, and this is expected to rise to 412 million by 2045^5^. Cardiovascular disease (CVD) deaths in Asia have nearly doubled from 5.6 million in 1990 to 10.8 million in 2019^6^. Addressing the rising burden of these major chronic disorders in Asian populations is a high priority for national and international stakeholders, including policymakers and healthcare providers.

Chronic diseases are complex and multi-factorial, and arise through the interaction of lifestyle, environment, and genomic factors^7^. Longitudinal population studies, in which people are characterised at baseline and followed up over time for health outcomes, play a unique role in identification of the proximal and upstream processes and aetiological mechanisms underlying chronic disease. However, existing prospective cohort studies with comprehensive phenotypic and particularly genotypic measurements are predominantly based on populations of European ancestry^8,9^. This not only represents an important global health inequity, but also a major opportunity for discoveries relevant to the health of the ∼4.8 billion Asian people living worldwide^10^.

Singapore, a city-state in Southeast Asia, is home to 5.6 million people, most of whom are of Chinese, Malay, or Indian ancestries. The presence of these three population groups living side-by-side, provides a unique opportunity to explore the diverse lifestyle and genetic profiles of people from East Asia, Southeast Asia, and South Asia, and to relate these to health trajectories. Endowed with highly advanced healthcare and research infrastructure, Singapore is ideally positioned to advance precision medicine and population health research, relevant to global Asian communities.

Here we describe the motivation, design, and early results of the Health for Life in Singapore (HELIOS) Study, a longitudinal population resource focussed on understanding the diseases and health states that are important to Asian populations. We show how the HELIOS study combines state-of-the-art clinical, molecular, and genetic epidemiological approaches, enriched with information derived from national health data, and highlight the extensive opportunities for transformative research. Our companion papers describe specific discoveries and innovations achieved using the study data, including findings directly relevant to health outcomes of people living in Asia^11–13^.

## Results

We recruited 10,004 Asian men and women aged 30 to 84 years to the HELIOS study between 2018 and 2022 (www.healthforlife.sg). Participants were recruited from the Singapore general population^11,12,14^. The cohort includes 6,784 people who identified as Chinese or other East Asian background, 1,807 people who are of Indian or other South Asian background, and 1,354 people of Malay or other South-East Asian heritage. There were 59 participants from other ethnicities (**Extended Table 1**). **Figure 1** briefly summarised our study design.

**Figure 1.**
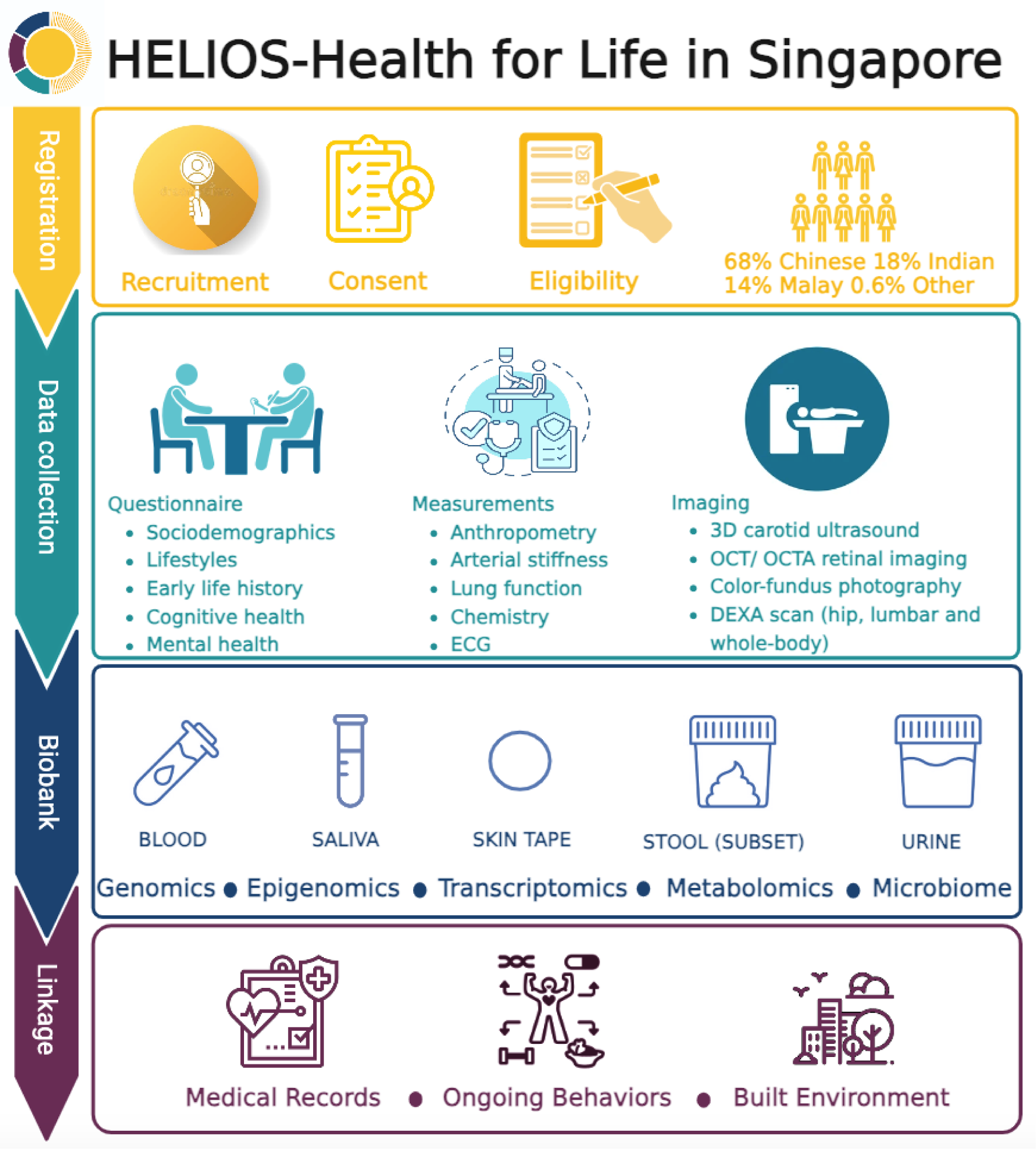
Overview of participant recruitment, data and biospecimen collection, and linkage. Abbreviations: DEXA: dual energy X-Ray absorptiometry; ECG: electrocardiogram; OCT: optical coherence tomography; OCTA: optical coherence tomography angiography.

### Disparities in health outcomes amongst Asian populations in Singapore

Despite similar age and sex distributions, our three Asian ethnic groups exhibit distinct profiles for health, including disease burden and distributions for clinically relevant exposures and endophenotypes. We highlight that Indian and Malay participants have a higher prevalence for hypertension, obesity, and type 2 diabetes (T2D), compared to Chinese participants (**Extended Figure 1**), and a higher frequency of symptoms for depression and anxiety. Waist circumference, waist-to-hip ratio, and visceral fat mass are also highest in Indian and Malay people. This is accompanied by increased levels of triglycerides, haemoglobin A1c (HbA1c), fasting plasma glucose, insulin, C-Reactive Protein (CRP) and other traits related to adiposity and insulin resistance (**Extended Table 1**). We also find evidence for differences in healthcare reach, across a wide range of actionable disease diagnoses. For example, compared to Chinese, Indians and Malays were more likely to have undiagnosed diabetes, while undiagnosed osteoporosis was common in all ethnic groups. These illustrations provide insights into the potential health gains that might be achieved through improved uptake and reach of healthcare interventions in our multiethnic Asian population (**Extended Figure 1**).

### Behavioural and upstream exposures relevant to chronic disease in Asian populations

Our study design enables exploration of ‘upstream’ behavioural, environmental, and social factors relevant to health in Asian communities. As an initial illustration, we show that density of food related amenities and the ratio of public to private housing correlate closely with the prevalence of diabetes in our cohort, a key exemplar of major chronic disease risk in the population (r=0.5 and 0.8, respectively; P<0.05, **Extended Figure 2**). Self-reported food intake of study participants shows divergent consumption of food items, nutrient composition, and differences in diet quality indices between the ethnic groups, that align closely with traditional Asian dietary habits (**Figure 2a-d**). While diet quality scores are associated with multiple cardiovascular and metabolic phenotypes within our Asian population groups, we highlight that dietary habit does not fully explain the differences between the ethnic groups. For example, while ‘favourable’ DASH (dietary approaches to stop hypertension) dietary quality score is highest amongst Indians, this directly contrasts with their high rates of obesity and adverse metabolic profiles compared to Chinese participants (P<2×10^-6^). Similarly, both self-reported and accelerometer-based objective measurements identify that total physical activity is higher, amongst Indians and Malays, despite their unfavourable patterns of adiposity and metabolic performance (**Figure 2e-g, Table 2,** P<2.2×10^-16^). Our data thus reveal striking variation between communities in behavioral, environmental, and social factors important for health. Interestingly, while our results confirm expected relationships with key clinical traits *within* ethnic groups, they do not fully explain health differences *between* populations. Our observations provide a strong motivation for deeper clinical and molecular epidemiological research focused on Asian populations.

**Figure 2.**
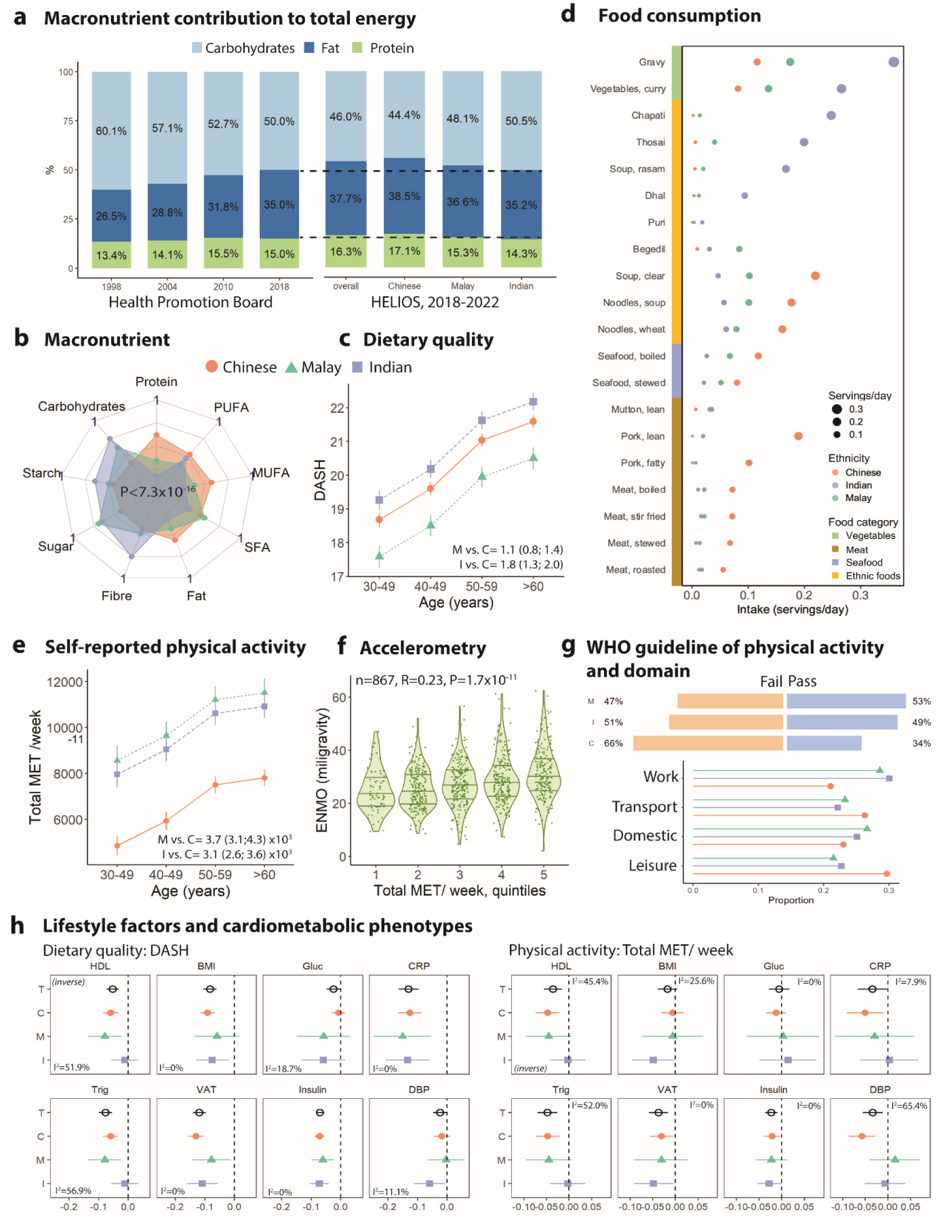
Lifestyle factors across three populations. **a)** Longitudinal changes in macronutrient trends nationally from 1998 to inception of HELIOS study in 2022 (n=10,004). Ethnic variations in **b)** macronutrients (proportion to total energy intake), **c)** diet quality represented by a modified DASH score (range from 0 for low quality to 35 for high quality) and **d)** top 20 FFQ foods (servings/day) significantly different across ethnicities within HELIOS study. **e)** Physical activity and **f)** Accelerometer-based physical activity according to the levels of self-reported physical activity (R=0.23, p=1.7×10^-11^). **g)** The proportion of people who meet the WHO guideline of physical activity by ethnicity and the proportion of physical activity across. **h)** The relationships between lifestyle factors and cardiometabolic phenotypes are heterogeneous across ethnic groups. **Abbreviations:** DASH: dietary approaches to stop hypertension; IPAQ: International Physical Activity Questionnaire; MET: metabolic equivalent of task; MUFA: monounsaturated fat; PUFA: polyunsaturated fat; SFA: saturated fat; WHO: World Health Organization.

### DNA sequence variation and functional genomic diversity

The disparate clinical profiles across the three ethnic groups are mirrored by extensive and structured covariation in molecular genotypes. Whole-genome sequencing (30x depth) reveals 252 million variants, including 239.7 million autosomal variants with 206.1 million Single Nucleotide Polymorphisms (SNPs) and 33.6 million short indels (**Figure 3a)**. Principal Component Analysis (PCA) and admixture analysis helped identify and cluster the dataset into three distinct population clusters corresponding to people of Chinese, Indian or Malay ancestry, as well as individuals who were admixed (**Figure 3b, Supplementary Figure 1**). The majority of variants identified are rare (minor allele frequency, MAF <1%; 95% and 90% of autosomal SNPs and short indels respectively), while greater than 50% of variants were observed only in one of the populations. Functional annotation using Annovar^15^ identifies 88,995 coding variants anticipated to impact protein structure (**Supplementary Table 1**). Among the coding variants, 6,130 are non-synonymous SNPs, 34 are protein truncating SNPs, and 82,831 are indels (**Figure 3a)**.

**Figure 3.**
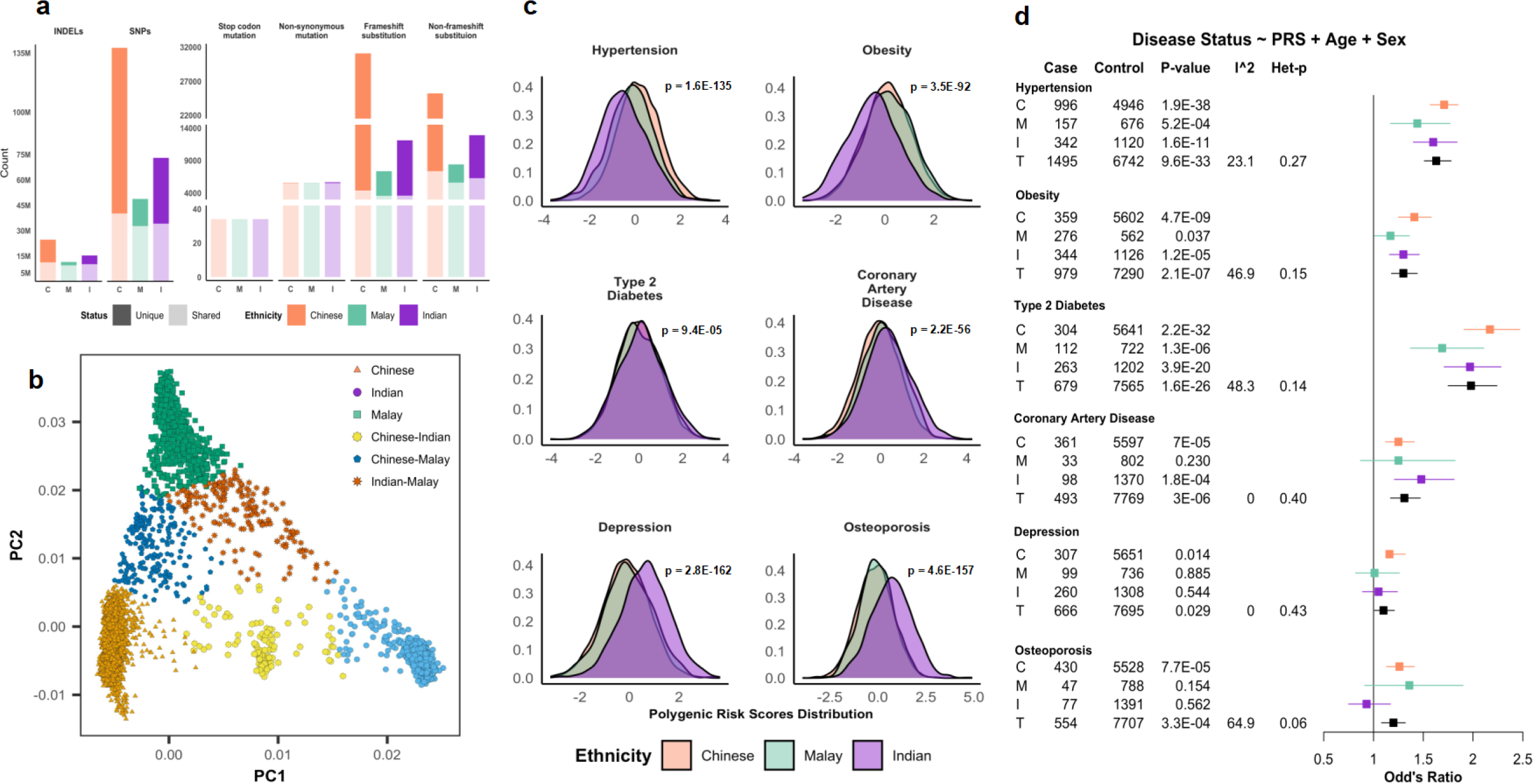
Genomic variation in Asian Populations. **a)** Number of variants annotated in each functional coding mutation category, by ancestry. The darker shades indicate unique variants observed only in the specific ancestry. **b)** 2-dimensional PCA genomic variants by ancestry group. **c)** Distribution of PRS scores for six complex traits in the three major ancestry groups. **d)** Forest plot displaying association PRS scores with respective complex trait by ancestry group, and overall [C: Chinese, M: Malay, I: Indian, T: Trans-ancestry].

Polygenic risk scores (PRS) confirm a strong relationship of genetic variation with quantitative traits and complex diseases, overall and in each of the three Asian ethnic groups. PRS also vary between populations (ANOVA p = 9.4×10^-5^ to 2.8×10^-162^; **Figure 3c**). The strongest separation was observed for depression, with higher PRS amongst Indians compared to Chinese and Malays (P=2.8×10^-162^). In contrast, although T2D PRS was strongly associated with diabetes risk amongst our Asian participants (OR for T2D 1.8 to 2.1 per SD, **Figure 3d**), T2D PRS shows limited variation between the Asian ethnic groups. Genetic factors identified by current Eurocentric genome-wide association studies thus also do not explain the three-fold higher risk of T2D observed amongst Indian and Malay individuals, compared to people of Chinese ancestry.

Quantification of DNA methylation in genomic DNA from whole blood (N=837,722 CpG sites), as a marker of genomic regulation reveals 16,444 unique CpG sites that are highly differentiated between the three Asian ethnic groups (P<2.9×10^-8^). These population specific methylation disturbances are enriched for location in DNase hypersensitivity sites (DHS), histone marks, enhancer and promoter regions, indicating ethnic specific patterns of genome regulation (**Extended Figure 3a, Supplementary Table 2**). The population-stratified methylation markers are enriched for location in the binding sites for specific, documented transcription factor across multiple cell lines (**Extended Figure 3b, Supplementary Table 3**). These include Pleiomorphic Adenoma Gene 1 (*PLAG1*) and Eleven-Nineteen Lysine-Rich Leukaemia Protein (*ELL*) (both P<10^-4^). PLAG1 is a nuclear transcription factor subject to maternal imprinting^16^, and which is implicated in pancreatic genesis, insulin secretion, and diabetes in neonates and adult organisms. We also note that the ethnically divergent methylation patterns strongly overlap CpG sites that predict future diabetes, providing evidence for nuclear regulatory disturbances that may contribute to the divergent metabolic outcomes observed between ethnic groups (**Extended Figure 3c, Supplementary Table 4**).

We used PCA to explore potential processes driving genome regulation in the population. We show that the perturbations in DNA methylation are enriched for association with educational attainment, dietary quality, adiposity and cardiometabolic health, based on directly measured and genetically inferred exposures (**Figure 4** and **Extended Figure 4, Supplementary Table 5, Supplementary Table 6**). Our results thus shine new light on the fundamental roles that these key modifiable social, behavioural, and physiological factors play, as primary, interlinked drivers of genomic regulation and health outcomes in diverse human populations.

**Figure 4.**
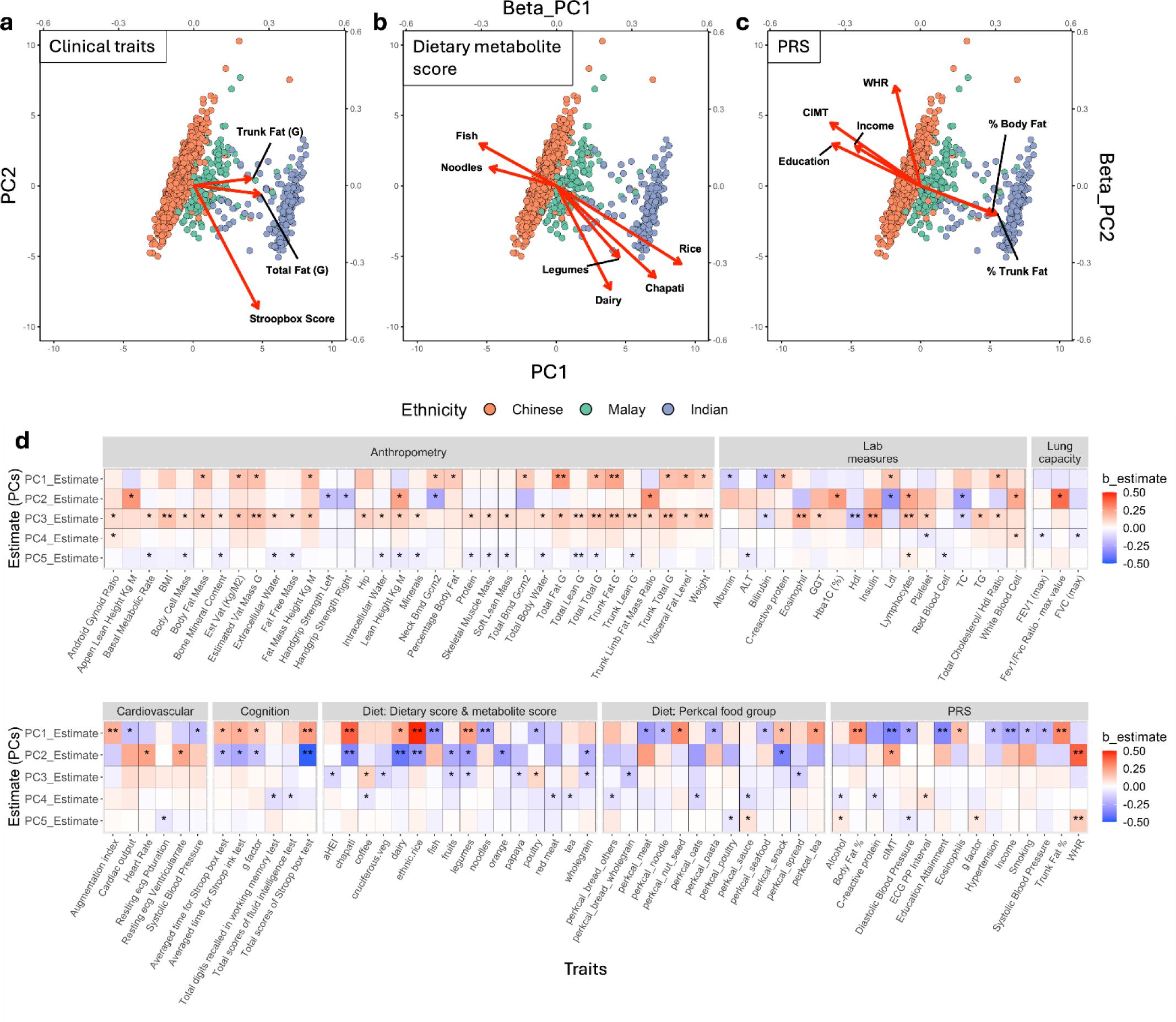
PCA plots of DNA methylation levels at 16,444 CpG sites that are highly stratified between our Chinese, Indian and Malay Asians and their association with various traits. **a-c)** display the methylation diversity for individuals from the three ethnic groups (PC1 on x axis, and PC2 on y axis). Overlaid are the effect sizes and directions for the beta coefficients derived from regression analyses of measured exposures on PC1 and PC2 of the DNA methylation; **a)** clinical traits; **b)** dietary exposures assessed objectively by circulating metabolites; **c)** Polygenic Risk Scores (PRS). The beta weights for PC1 and PC2 are scaled along the top x-axis and the right y-axis respectively. The results identify the directly measured and genetically inferred exposures that may relate to population level epigenetic variation between Asian ethnic groups. **d)** The effect sizes and the directions for the beta coefficients derived from regression analysis of measured exposures on PC1 to PC5(%Variance base on top 100 variance; PC1 – 17.6%, PC2 – 9.9%, PC3 – 5.1%, PC4 – 2.2%, PC5 – 2%). *p-value<0.05, **p-value<0.0015. Abbreviations: CIMT: carotid intima-media thickness; WHR: waist hip ratio.

### Metabolic variation in Asian populations

Metabolomic profiling of plasma by high-throughput semi-quantitative mass spectrometry enabled us to quantify plasma concentrations of 1,073 discrete metabolites. We show that dietary patterns of our Asian participants intersect closely with their metabolic variation, enabling identification of metabolite sets that are representative of Asian dietary patterns; these associate closely with perturbations in regulatory pathways, and predict multiple chronic diseases.^12^ We also find that 153 of the 1,073 plasma metabolites characterized show marked divergence between all three Asian ethnic groups (P<1×10^-5^, **Figure 5a, Supplementary Table 7**); of these 128 metabolites are of known identity. In general, amongst the 153 highly differentiated metabolites, Indians and Malays had lower levels of lipid metabolites, and higher levels of amino acids and nucleotides compared to Chinese (**Figure 5b, Supplementary Table 7, Supplementary Table 8**). 63% of lipid metabolites were inversely associated with the presence of hypertension, obesity, T2D, or CVD (P<4.7×10^-5^), and 16% were inversely associated with all four phenotypes. Age, sex, genetic ancestry, diet, and BMI were each determinants of plasma concentrations for the highly differentiated metabolites (**Figure 5c**), but with substantial differences in their contribution on a metabolite specific basis. For example, BMI accounted for 18% of the variation in glutamate, while age accounted for 20% of the variation in the androgenic steroid dehydroepiandrosterone sulphate. Strong effects for genetic ancestry on metabolic variation were seen for 1-margaroyl-2-arachidonoyl-GPC (17:0/20:4), a phosphatidylcholine derived from eggs, fish, and meat.^17–19^ We show that concentrations of this metabolite are positively associated with self-reported intakes of red meat (P=5.3×10^-57^), fish (P=6.5×10^-38^), dairy (P=1.4×10^-20^), and poultry (P=1.1×10^-17^). Levels are also associated with chapati consumption, that is common in Indian communities (P=1.6×10^-3^). Circulating 1-margaroyl-2-arachidonoyl-GPC levels are strongly influenced by genetic variants in the *FADS1*/*FADS2* gene cluster, a highly pleiotropic region that is linked to multiple lipids, cardiometabolic, inflammatory traits, skin diseases and pregnancy outcomes.^20^ *FADS1/FADS2* variants are also known to be stratified between Asian populations, and recognised to influence metabolic responses to dietary intake, and may provide the basis for genomically determined ‘Precision Nutrition’.^20^ Our observations further highlight the important roles for both genetic and lifestyle factors in driving divergent metabolite profiles and health outcomes amongst Asian people.

**Figure 5.**
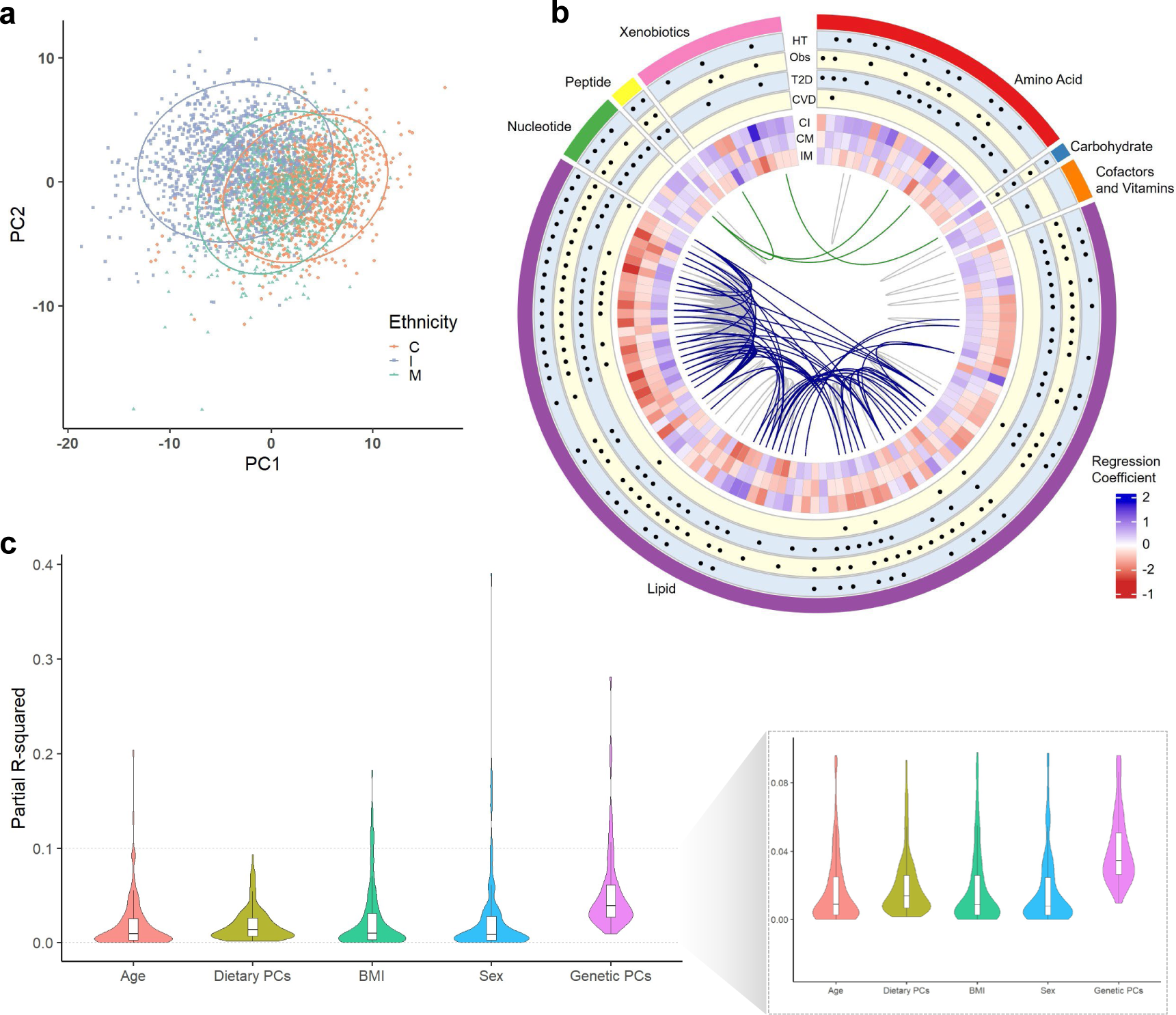
Metabolic variation across three populations. **a)** PCA plot of 153 significantly differentiated metabolites across three age-sex matched ethnic cohorts of 1146 individuals each (The first two PCs explain 23% of variation). Selection criteria for 153 metabolites include: 1) significantly associated with ethnicity in the discovery cohort (70% participants, P<1×10^-5^), and 2) significantly associated with ethnicity in the test cohort (30% participants, P<0.05 and same direction of estimates as in the discovery cohort). **b)** Circos plot of 128 well-characterized and known metabolites (in sequence from outermost to innermost layer): 1) metabolite super-pathways, 2) significant associations with HT, Obs, T2D, and CVD, denoted by a black dot, 3) estimates of regression coefficient for association with ethnicities (CI: Indian compared to Chinese, CM: Malay compared to Chinese, IM: Malay compared to Indian). Curved lines at the centre highlight significant pairwise correlation between metabolites. Grey lines represent pairwise correlations within the same super-pathway; blue lines represent pairwise correlations across sub-pathways but within the same super-pathway; green lines represent pairwise correlations across super-pathways. **c)** Violin plot showing contribution (as partial r-squared values) of age, dietary PCs, BMI, sex, and genetic PCs on variation of plasma abundance of 153 metabolites. The inset plot zooms in on the partial r-squared distribution between 0.0 - 0.1. Abbreviations: BMI: body mass index; CVD: cardiovascular disease; HT: hypertension, Obs: Obesity, PCA: Principal Component Analysis; T2D: type 2 diabetes.

### Potential for discovery through molecular epidemiological studies of Asian populations

The clinical, molecular, behavioural, and environmental diversity between the Asian ethnic groups provides robust new opportunities for discovery relevant to human biology and health outcomes. To illustrate this, we carried out genome-wide association of the 153 ethnically diverse plasma metabolites. We identify 365 independent genetic variants in 140 genomic loci, that are significantly associated with 113 metabolites at a genome wide significance threshold (P< 5×10^-8^) (**Figure 6a**). We observe a strong degree of genetic pleiotropy at multiple loci, in particular the *FADS1/FAD2* gene locus which was associated with 39 metabolites **(Figure 6b)**. Summary-data-based Mendelian Randomisation (SMR) analysis of metabolites with cis-eQTLs identified 1,176 significant gene-metabolite pairs after multiple testing correction (P<4×10^-5^), comprising 585 genes and 104 metabolites **(Supplementary Table 9)**. We were able to replicate 166 gene-metabolite associations and identify 51 additional associations using cis eQTL information obtained using the HELIOS transcriptomics data (**Supplementary Table 9, Supplementary Table 10**). Colocalization analysis reveals shows that 79 of these gene-metabolite pairs are likely to share a common causal variant (coloc-H4 P>0.7). This includes the novel finding that plasma concentrations of dopamine 3-O-sulfate, are influenced by genetic variants at the *cis*-eQTL locus for *SMAD5,* a transcriptional regulator protein involved in the TGF-Beta pathway (**Figure 6c**) and implicated in the development of dopaminergic neurones.^21^ Similarly, variation in plasma levels of metabolite X-11381, are determined by genetic variation found at the cis-eQTL locus for *Nephrocystin 4* (*NPHP4*, **Figure 6d),** which plays an important role in renal tubular development and function. X-11381 is also associated with raised blood pressure and cardiovascular disease in our cohort (false discovery rate, FDR– P<0.05; **Supplementary table 7**). Our rich multi-omics data thus provide multiple opportunities to improve understanding of the molecular pathways influencing metabolic performance and other pathways leading to chronic disease in Asian populations.

**Figure 6.**
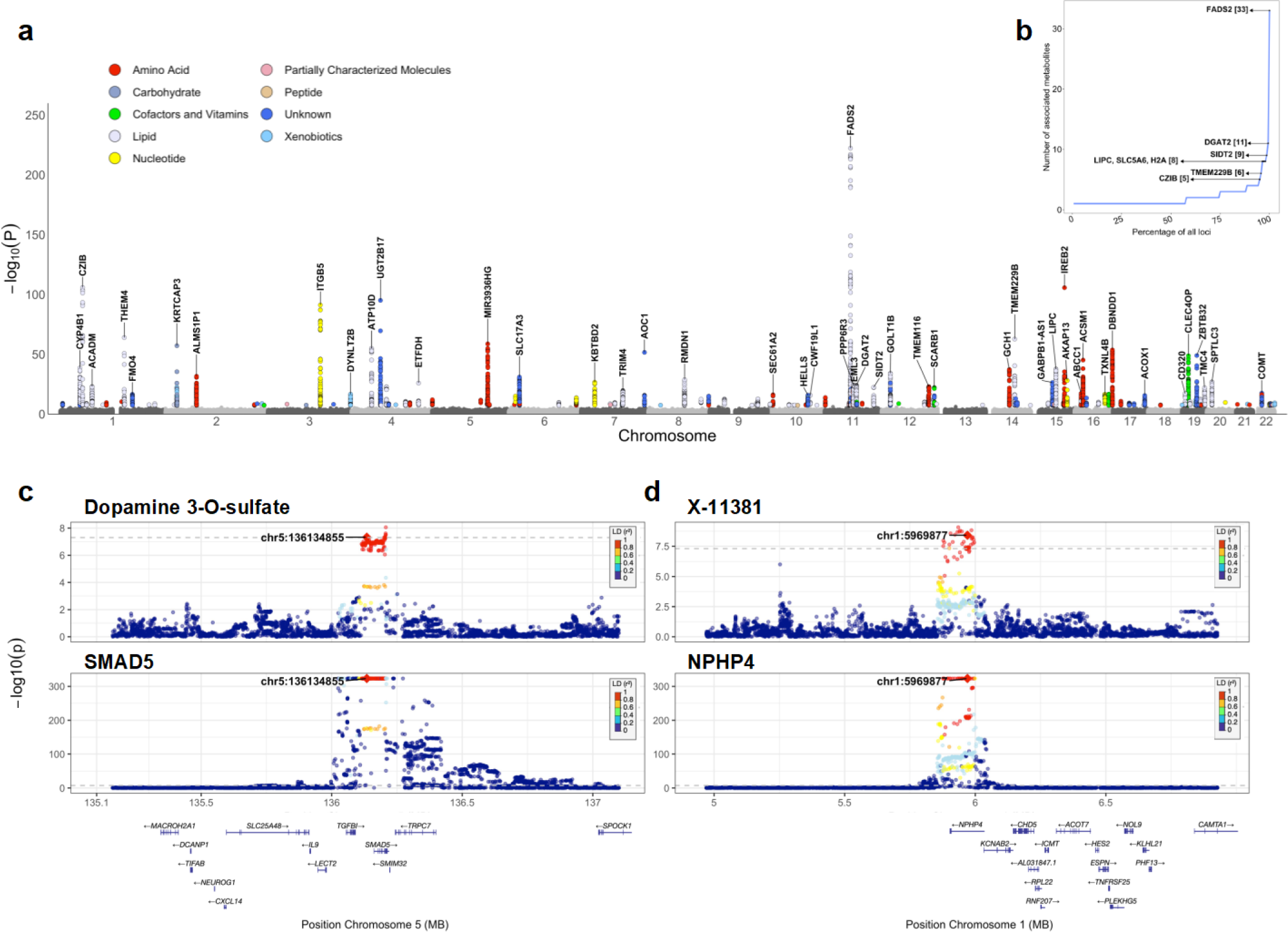
Genetic Architecture of Molecular Traits. **a)** Manhattan plot for summary of all associations between plasma metabolite levels and genetic loci. Only genetic variants with P<5×10-8 are coloured based on the strongest associated metabolite group for the specific variant. Genes for top 50 loci identified through SMR are annotated. **b)** Distribution of number of associated metabolites per locus, demonstrating the pleiotropy of genetic effects on metabolites. The loci with at least 5 associated metabolites are annotated with the SMR associated gene. **c)** Regional plot highlighting the shared causal variant between Dopamine 3-O-sulfate and SMAD5. **d)** Regional plot highlighting the shared causal variant between X-11381 and NPHP4.

### Linkage to national health and administrative records

With participant consent, we link HELIOS research phenotypic data securely to their national health data, using the NRIC, a unique national identifier that is held by Singaporean citizens and Permanent Residents. De-identified linked research, health and administrative data were made available through the Trusted Research and Real-World Data Utilisation and Sharing Tech (TRUST) platform (https://trustplatform.sg). National Health and Administrative Records were identified for 95% of study participants, and include national disease registry records, disease diagnosis, national insurance claims, medications, laboratory tests, radiology, surgical procedures, and death registry records from 1998 to 2020. The linked national health and administrative records for our 10,004 participants include 1.6 million laboratory test results, 776,505 prescriptions and 131,211 diagnostic episode codes. Using diabetes as a case study, we show that the national health data recapitulate age stratified, ethnic disparity disease risk, and enable identification of incident diabetes cases, with greatest risk amongst participants who are older, obese and impaired fasting glucose **(Extended Figure 5**). These linked national data thus provide deep opportunities to extend baseline health assessment of participants, and to identify future health trajectories, including incident disease.

### Reproducibility of measurements

We demonstrate the reproducibility of our research phenotypic characterisation, by carrying out repeat assessment of 398 participants, one year after enrolment (range 58 to 1073 days). We show moderate to strong intra-class correlations for measures made, in all domains of assessment, a performance that is similar or better than those reported by UK Biobank^22^ and other major population studies.^23^ In general, objective physiological measurements were more reproducible than self-reported lifestyle and cognitive measurements (**Extended Figure 6**). High data completeness and reproducibility further support the validity of our unique multiethnic Asian dataset.

## Discussion

Asian populations are widely recognised to be under-represented in global genomic and health-related research cohorts, compared to their European counterparts. This represents an important impediment to identification of the population specific behavioural, environmental, genomic, and molecular exposures and processes that impact Asian health. The limited ethnic diversity of existing population studies also represents a major obstacle to the development of effective and evidence-based approaches for accurate diagnosis and therapeutic intervention, that address the health needs of Asians.

To advance beyond current state-of-the-art, we have established the HELIOS study, a deeply phenotyped, longitudinal population cohort comprising 10,004 men and women from the multi-ethnic Asian population of Singapore. Our participants underwent extensive clinical, behavioural, environmental, and molecular characterisation, adopting techniques that are validated, aligned to best practices, and directly interoperable with international precision medicine cohort studies. HELIOS includes people of Chinese (East Asian), Malay (Southeast Asian) and Indian (South Asian) background. The inclusion of these three major Asian ethnic groups provides an opportunity for precision medicine research, that has the potential for relevance beyond Singapore, and across the wider Asia-Pacific region. The clinical characteristics of the cohort are broadly representative of the population from which they were recruited and are notable for the high rates of diabetes and related metabolic disturbances, that are recognised to be highly prevalent amongst Asian people.

Whole genome sequencing demonstrates the genetic diversity of the population. While the majority of individuals cluster in one of the three main ancestral groups, there is also evidence for recent population admixture between each of these three groups. This unique population genetic architecture provides the basis for the presence of functionally and clinically relevant DNA sequence variation, that is specific to Asian subgroups. Characterisation and interpretation of this population genetic variation is anticipated to provide opportunities for new discoveries relevant to disease aetiology and is also an essential prerequisite for the application of genomic medicine in Asian populations.

Our comprehensive characterisation of study participants is specifically designed to capture a wide spectrum of exposures relevant to health, as well as to reveal the systems biology that links these exposures to phenotypic variation and health outcomes in Asia. In keeping with this approach, we demonstrate the presence of variation in genome regulation and metabolic performance between the three Asian ethnic groups. For example, we identify extensive, ethnic-specific perturbations in DNA methylation that intersect with PLAG1, a nuclear transcription factor linked to pancreatic biology and diabetes,^24^ which overlay CpG sites are linked with obesity and diabetes,^25^ mirroring the divergent metabolic outcomes between ethnic groups. Stratified genomic regulation correlates with socio-economic factors, population specific dietary habits, physical activity, adiposity, and genetic diversity; these observations shine new light on the fundamental roles that social, behavioural, and inherited factors play as interlinked drivers of health outcomes in diverse human populations. We further use metabolomic profiling to demonstrate extensive variation in metabolite concentrations between our Asian groups, that reflects their specific patterns of diet, the differing levels of adiposity, the presence of genetic variation and transcriptional control. In parallel, metabolite profiling of our unique Asian population cohort has enabled identification of previously unrecognised pathways underlying cholesterol transport and cardiovascular risk, and potential opportunities for novel therapeutic approaches to cardiovascular disease prevention.^13^

Linkage to national health data relevant to health provides powerful opportunities to enrich baseline phenotyping of participants in population-based cohorts, as well as to identify longitudinal health outcomes efficiently and accurately. Longitudinal population cohorts in Europe and North America have a long tradition of successful record linkage that has accelerated health-related research in these settings. In contrast, record linkage has been uncommon amongst the available Asian population cohorts, reflecting both limited implementation of national health data, as well as the nascent state of regulatory frameworks to enable safe data integration. Here we demonstrate the ability to achieve linkage to health and administrative records amongst the multi-ethnic Asian populations of Singapore, using a secure platform for linkage, deidentification and analysis, hosted by the Singapore Ministry of Health (the TRUST platform). We use this framework to retrieve extensive medication, laboratory, and diagnostic data for our study participants. With diabetes as a use case, we demonstrated the ability to identify accurately people with diabetes both cross-sectionally and prospectively, and to show expected longitudinal risk relationships. This successful approach to secure record linkage is unrivalled in the Asia Pacific region and will be instrumental in advancing the research goals of the study, for the benefit of Asian people living in Singapore and other global settings.

With rich, multi-layered baseline data and long-term follow-up through linkage, the HELIOS study provides a world class resource for biomedical researchers from a wide range of disciplines, to investigate the behavioural, environmental, genomic, and molecular factors impacting health in Asian populations, with a level of detail that has not been previously possible. The successful approaches to population-based research established in the HELIOS study also provides the blueprint for ongoing efforts to create a precision medicine cohort comprising 100,000 people, the SG100K population study, to enable national efforts to advance precision medicine for Asian populations.

## Methods

HELIOS is a prospective population-based cohort, comprising men and women aged 30 to 84 years, living in Singapore (www.healthforlife.sg; https://www.instagram.com/heliossg100k/; https://www.facebook.com/HELIOSSG100K/, IRB approval by Nanyang Technological University: IRB-2016-11-030). Study design was informed by initial pilot studies (N=184, recruited between January 3, 2018 and March 21, 2018, **Supplementary Table 11**), which enabled development of community engagement and involvement activities, study protocols and training programs. Participants were recruited between April 2, 2018 and January 7, 2022. Assessment of reproducibility was carried out in 398 participants (recalled between September 3, 2019 and January 28, 2022, **Supplementary Table 12, Extended Figure 6**). The study is a template for future efforts with increased sample size (SG100K, target 100,000 participants).

- *Recruitment*. Study participants were recruited from the Singapore general population through a range of community outreach programs to ensure participation from ethnic minority groups, as well as people across socio-economic groups. Community engagement included language-specific recruitment drives conducted in the worship places, religious associations, and community associations across Singapore; multilingual study advertisement and documents (English, Chinese, Malay, and Tamil); and collaboration with a range of employers and occupational groups. Individuals were excluded if they were pregnant or breastfeeding, or had acute illness, major surgery within the previous 3 months, current participation in a drug trial, or cancer treatment in the past year.
- *Consent.* HELIOS asks permission from participants to use the data and samples that they contribute, for clinical and molecular epidemiological research focussed on improving human health. This includes the application of ‘untargeted’ molecular profiling techniques that assess genomic, proteomic, transcriptional, metabolomic and other ‘omic’ variation in the biological samples collected. Participant consent also includes permission for linkage to disease registers, medical records, social care datasets and other health-related datasets held by Singapore’s public bodies. Linkage is enabled by the Singapore NRIC, a unique national identifier allocated at birth, and with universal coverage. Consent provides permission for use of the data and samples from participants, by both academic and industry researchers, and for recontact of participants, including recontact based on phenotypic or genotypic characteristics. The HELIOS study operates under the governance framework of the Nanyang Technological University, and with Institutional Review Board approval (Ref: 2016-11-030)
- *Baseline examination.* At enrolment, participants complete a comprehensive physiological, clinical, and behavioural assessment, carried out in a single visit (**Extended Table 2**). The electronic health and lifestyle questionnaires collect demographic, lifestyle, reproductive history, and other potentially health-related information. In addition, a broad range of physiological measurements, including a state-of-the-art imaging module comprised of a 3-D carotid ultrasound, dual energy X-ray absorptiometry (DEXA) scans for bone density and body composition, and comprehensive optical imaging, are performed. These imaging technologies will enable the identification of pre-clinical disease phenotypes that will aid prognostic and preventative research. Participants also complete a physical fitness test and have physical activity monitored using accelerometer devices over a 7-day period. Biological samples (blood, urine, saliva, stool, and skin tapes) are also collected. The assessment process, biological samples collection and storage, quality management, return of assessment findings, ethics and data security are described in detail in **Supplementary Methods**.
- *Follow-up.* The HELIOS study will follow up participants long-term to identify any event of interest. This design allows the investigation of the causes and nature history of a broad range of diseases and health conditions. Participants enrolled in the HELIOS study will be followed up through routine health record linkage, re-contact with participants, Singapore Cancer Registry, Ministry of Health, and Health Promotion Board records, where available, for medical records, ongoing behaviours and built environment exposure.

### Analysis of biological samples

- *Clinical chemistry*. This includes assessment of fasting glucose, insulin, and lipid profile, as well as HbA1c and CRP. Fasting glucose, HbA1c and lipid profile were measured from fasting blood samples by the accredited laboratory (QuestLab, Singapore, SAC–SINGLAS ISO 15189:2012). Fasting insulin and CRP were measured with immunoassays using the ADVIA Centaur XPT Immunoassay System and ADVIA 1800 Chemistry System, respectively (Siemens Healthcare, Erlangen, Germany).
- *Whole Genome sequencing (WGS)*. Whole genome sequencing was carried using the Novaseq platform, with data processing using DRAGEN v3.7.8. Individual sample Variant Call Format (VCF) files were transformed into HAIL matrix tables^26^. Multi-allelic sites were efficiently split into multiple rows of bi-allelic sites, ensuring a comprehensive representation of the genetic variation. Samples were merged in batches of 1,000, to create a unified HAIL matrix table representing the sample cohort, with 258,062,302 genetic variants. Stringent variant and sample quality control (QC) parameters were employed to ensure the accuracy and reliability of the genomic data. These included a number of q30_bases (threshold 77.5GB high quality bases), as well as ratios for transition/transversion, heterozygous/homozygous variation, and insertion/deletion, applying a threshold of 6x Median Absolute Deviation (MAD) for each. Samples exhibiting more than 1% cross-contamination, call rate <95%, autosomal coverage <95% at 15X, or discordant sex information (reported vs genetically determined) were also flagged. The QC metrics were added as annotations to the HAIL Matrix table, which was then converted to a merged VCF file of 10,000 samples. The VCF file was also converted and stored as PLINK2^27^ binary files to perform downstream analysis.
- *Methylation profiling*. Bisulfite conversion of genomic DNA was performed using the EZ DNA methylation kit (Zymo Research, Orange, CA), with DNA methylation quantified using the Illumina Infinium MethylationEPIC BeadChip® array (EPIC) (Illumina, Inc, CA, USA) according to manufacturer protocols. Bead intensity was retrieved using the *minfi* package in R, with a detection P<0.01 used for marker calling. Of the 846,604 positions assayed by the array, we excluded markers with call rates <95% (N=8,882). In total 58 samples were excluded, 2 for array scanning failure, 39 for sex inconsistency and 17 duplicates. None of the samples failed sample call rate (<95%). This left us with 837,722 CpG sites and 2,342 samples for analysis. We analysed epigenome-wide data in R using *minfi* and other R scripts, in accordance with the CPACOR pipeline.^28^ In brief, marker intensities were normalised by quantile normalisation, with white blood cell subsets imputed.^29^
- *RNAseq.* RNAseq libraries were prepared using samples of whole blood (n=1,234) collected in PaxGene RNA tubes at enrolment. RNAseq libraries were prepared from at least 1μg of total RNA using NEBNext® Ultra™ II Directional RNA Library Prep (New England Biolabs, Inc.), with GLOBINClear (Thermo Fisher Scientific) for depletion of globin gene RNA and Ribosomal RNA (rRNA). The libraries were sequenced on a NovaSeq6000, using a paired-end run of 2 x 150bp. We aimed for at least 30M aligned reads per library (∼9Gb of data). Adapter and quality trimming were performed in TrimGalore^30^ whereas SortMeRNA was used for the removal of rRNA.^31^ Alignment to the reference genome (GRCh38) was done using STAR version 2.7.9a^32^, followed by quantification of reads with RSEM version 1.3.3.^33^, which identified a total of 60,708 genes. Gender mismatch check was performed by interrogating for anomaly across 5 genes – namely XIST, RPS4Y1, EIF1AY, DDX3Y, and KDM5D. A total of 6 samples had failed this check, resulting in a total of 1,228 samples for downstream analysis. Genes with transcript per million (TPM) ≥1 and read count ≥6 in at least 20% of the samples were retained; resulting in a remaining total of 12,434 genes. Finally, the genes were normalized using the Trimmed Mean of the M-values (TMM) approach^34^.
- *Metabolite profiling*. The Metabolon Global Discovery Panel was used for untargeted mass-spectrometry-based metabolic profiling of 10,000 fasting EDTA plasma samples. Samples were initially stored at -80C, then thawed, aliquoted, and shipped on dry ice to Metabolon. Samples were prepared and extracted for assay using four methods: two separate reverse-phase (RP)/UPLC-MS/MS methods with positive ion mode electrospray ionization (ESI), RP/UPLC-MS/MS with negative ion mode ESI, HILIC/UPLC-MS/MS with negative ion mode ESI. All methods utilized a Waters ACQUITY ultra-performance liquid chromatography (UPLC) and a Thermo Scientific Q-Exactive high resolution/accurate mass spectrometer interfaced with a heated electrospray ionization (HESI-II) source and Orbitrap mass analyzer operated at 35,000 mass resolution. Several recovery and internal standards, and controls (blanks and pooled matrices) were added for quality control (QC) purposes. Experimental samples were randomized across the platform run with QC samples spaced evenly among the injections. Five samples failed Metabolon QC standards and were removed from analysis. Peak area-under-curve was used for metabolite quantification, and data across inter-day batches were normalized by median scaling. Data corresponding to (i) 235 samples from a second visit of the same participant, and (ii) 9 outlier samples (greater than 6 standard deviations from the mean of first and second principal components) were excluded. Metabolites missing in more than 25% of the data were removed, and the remaining imputed with minimum value, then log-transformed and standardized before further data analysis.

### Statistical analyses

- *Clinical definitions.* Hypertension was defined as self-reported and/or blood pressure ≥140/90 mmHg; obesity is BMI ≥30 kg/m^2^.^35^ Type 2 diabetes was self-reported and/or fasting glucose ≥7.0mmol/L or HbA1c ≥6.5%.^36^ Cardiovascular disease includes subclinical atherosclerosis, defined as the presence of atherosclerotic plaque or mean cIMT ≥0.8.^37^ Depressive symptoms are PHQ-9 score ≥10^38^, whereas anxiety symptoms: GAD-7 score ≥10.^39^ Osteoporosis is defined as lumbar spine bone mineral density T-score of -2.5 or below.^40^
- *Correlation across phenotypes*. The correlation coefficients across phenotypes were calculated using Pearson correlation analyses for z-scored transformed measurements and visualised in heatmap.
- *Dietary habit and nutrition*. Ethnic variations in dietary intakes (foods and macronutrients) and diet quality (DASH score)^41^ were assessed using the validated FFQ^42^. Food items were recorded as servings/day, and macronutrients were expressed in kcal/day after accounting for type of macronutrients/serving and weight/portion of each food, and subsequently aggregated to derive % contribution to total daily energy intake. *For macronutrients*, % macronutrient was scaled and visualised as radar plot (R package *fmsb*) across ethnicity. *For food items*, daily servings were log-transformed and analysed using linear regression, adjusted for age, sex and ethnicity, with Chinese being the reference. We applied Bonferroni-Hochberg corrected p-value threshold of P<1×10^-100^ and selected the top 20 foods significantly higher in Malay and in Indian subgroups. Foods were grouped into 4 categories based on animal source or relevance to ethnicity. *For DASH*, a modified score was derived from 7 components (fruits, vegetables, wholegrains, nuts and legumes, low-fat dairy, red and processed meats, and sweetened beverages), ranging from 0 (low quality) to 35 (high quality). Difference across ethnicity was analysed using linear regressions.
- *Physical activity*. Physical activity in various domains and intensity levels and sedentary behaviour were derived based on the validated long IPAQ^43^. Participants with at least 150-300 minutes of moderate-intensity or at least 75-150 minutes of vigorous-intensity physical activity or an equivalent combination of moderate- and vigorous-intensity activity throughout the week and with at least 2 or more days on moderate- and vigorous-intensity activities a week were deemed as meeting the WHO guidelines for recommended physical activity^44^. Accelerometry data were collected as an optional assessment amongst the first 1000 participants in the next phase of the HELIOS Study. Participants wore Axivity AX3 wrist-worn triaxial accelerometer on their non-dominant wrist continuously for 7 consecutive days, including during sleep. Raw accelerometry data were calibrated to local gravitational acceleration^45,46^ following which movement-related acceleration was expressed using the Euclidean Norm Minus One (ENMO) metric (https://github.com/MRC-Epid). This method has been validated against energy expenditure in free-living conditions^45,46^ to generate mean Euclidean Norm Minus One (ENMO). The data of 867 out of 1000 participants were suitable for analysis. Comparison across ethnicities were performed using Kruskal-Wallis rank sum and Chi-square tests.
- *Environment*. OneMap APIs (https://onemap.gov.sg) were called within the R environment to generate the latitude and longitude of each participant’s postal code and planning area. All geospatial Singapore data with relevant attribute tables were extracted from the national open data collection (https://beta.data.gov.sg). The extracted tables include planning area, population census by subzones; subzones by type of dwelling; and parks and nature reserves. Open-sourced QGIS v3.32.1 software was used to project geospatial data and population density. Geospatial tags for shops selling food and beverages and shopping centres, as well as amenities for sustenance, were extracted using QuickOpenStreetMap plug-ins. OpenStreetMap IDs representing food amenities (n=9901) were tagged to the respective planning area in Singapore using OneMap API. To generate bubble plots linking environmental factors with disease outcome, the area (m^2^) per planning area polygons was calculated to derive population density using *sf* and *lwgeom* package.
- *Annotation of Genetic Variants*. Variants were annotated using the ANNOVAR tool^15^, with the refGene GRCh38 reference. Novel variants present in our dataset were identified after comparing the dbSNP v156 database^47^ of all reported variants.
- *Population Structure analysis and clustering*. To understand the genetic structure and stratify our population, we applied strict filters to the data excluding variants with MAF < 5% (i.e. present in less than ∼500 out of the 10,000 genotyped individuals), Hardy-Weinberg equilibrium (HWE) P<0.1%, sample and variant missingness <2% and removed variants in the MHC region as well as the Chr8 Inversion region. Duplicated samples (n=3) and samples with reported ancestry labelled as “Others” (non-Southeast Asian [n= 60]) were removed for the current analysis. Linkage Disequilibrium (LD) based pruning was performed for the final filtered data with an LD-r^2^ of 0.1 within a 200KB window. Genetic relatedness was estimated using the genome function to determine the pi_hat estimate for all pairs of individuals in our dataset. We performed PCA to extract the top 50 genetic PCs from our data. Given the complex structure of our data, we use a data-driven approach to determine and cluster the individuals belonging to specific ethnic groups. The results of the PCA analysis were used to perform K-means clustering (K=3) to group the individuals into three super populations (Chinese, Indian and Malay). These ancestry labels were used to estimate the ancestry-stratified allele frequency file, which is used as input to run supervised admixture analysis using SCOPE.^48^ The results from the admixture analysis were used to determine the 6 final ancestry clusters using a semi-supervised K-means clustering approach. Additionally, to understand the genetic structure of our data with reference to 1000 genomes^49^, we merge the LD independent SNPs with 1KG data from four super populations and perform PCA again with the merged set of samples and variants after applying the same filters as above. All the filtering, PCA, LD and relatedness analysis were performed using the PLINK2 tool^27^ and the k-means clustering was done using R.
- *PRS*. Summary statistics for estimating PRS for the genomic and the epigenomic variation analysis were obtained from the PGS Catalog (**Supplementary Table 13)**^50^, selecting the study with best possible trans-ancestry base data and validation. PRS was estimated using the score function in PLINK2^27^, separately for each ancestry group, and then merged and normalized to identify ancestry level differences. The performance of PRS was tested separately for each ethnic group while adjusting for age and sex, and meta-analysed to determine trans-ancestry performance. For the PRS used in the methylation analysis, scores were estimated together for all the individuals with methylation data available.
- *DNA methylation.* We first identified CpG sites that were considered significantly differentially methylated between any pair of Asian ethnic subgroups at a p-value threshold of 2.9×10^-8^. This cutoff was obtained via a two-step process. Firstly, we defined epigenome-wide significance as P<8.62×10^-8^, which was obtained via permutation testing and is also close to what would have been obtained via Bonferroni correction. We then performed a second Bonferroni adjustment for the multiple testing between the three pairs of ethnic subgroups (Chinese versus Malay, Chinese versus Indian, and Malay versus Indian), which brings us to P<2.9×10^-8^. To further assess the relationship between DNA methylation and metabolic outcomes, we focused on 315 sentinel CpG sites that are significantly associated with incident T2D based on our epigenome-wide association testing performed in age-, sex- and ethnicity-matched controls in the Translating Omics into A Stratified approach for prevention of T2D (TOAST) study. As one of the CpG sites was not found in HELIOS, this left us with 314 CpG sites for the analyses. DNA methylation was measured using baseline samples collected before onset of T2D, with primary analysis of epigenome-wide data performed as described previously^28^. In brief, the association of each autosomal CpG site with incident T2D was tested using logistic regression, adjusted for confounders such as age, sex and further adjusted for imputed white blood cells (WBC) proportion and PC1-30 of control probe intensities. To assess the association of these CpG loci with BMI in the HELIOS participants, we then performed linear regression with the same covariate adjustments. Correlation between CpG sites were assessed using Pearson correlation analyses, with the circos plot generated by the *circlize* package.
- *Functional Annotation of Sentinel CpG:* We perform functional overlap analysis and annotation of the sentinel CpGs using eFORGEv2.0^51^ analyzing the 16444 CpG sites for enrichment across DNase I hotspots, 5 histone marks and 15 chromatin states across 39 cell types from the Roadmap Epigenomic Consortium.^52^ We determine the number of Sentinel CpGs overlapping with the annotated regulatory and chromatin regions in the different cell types. The enrichment of our sentinel CpG set was evaluated by comparing it to 1,000 background sets that contain an equal number of sites as the input. The background sets were matched using gene annotation and CpG island annotation and the mean overlap for the background sets was calculated. We used the background sets to calculate the fold enrichment as observed count /mean (expected counts) and obtained an empirical P value from the distribution of the background sets.
- *Transcription Factor (TF) Enrichment:* The binding site information for the 1210 human TFs tested was obtained from the Remap database, 2022 release (https://remap.univ-amu.fr/).^53^ We used the homo sapiens Cis Regulatory Modules (CRM) peaks for this analysis. We first determine how many of our sentinel CpGs overlap with the binding sites of the different TFs, and then estimated the fold enrichment; p-value for enrichment was calculated by comparing the overlap of our sentinels to the overlap of CpG probes from the background set of all CpGs. The p-value for enrichment was obtained using hypergeometric test and corrected for multiple testing at a False Discovery Rate threshold of 0.05 (**Supplementary Table 4**).
- *Enrichment across behavioural, lifestyle and genetically inferred traits.* We tested the associations between the 16,444 ethnically differentiated CpGs, and 187 trait-exposures, including directly measured phenotypes as well as PRS to derive genetically inferred exposures (**Supplementary Table 5**). Linear or logistic regression was used, with adjustment for age, sex, ethnicity, methylation array control probe PCs, and white cell subset composition estimated by the Houseman method^29^. We then performed the same analysis for all CpGs on the MethylEPIC array (837,722) to estimate background expectations. We then calculated enrichment (observed vs. background), using the hypergeometric test. We inferred statistical significance at P<0.05/32, based on an estimate of 32 independent phenotypes derived from PCA of phenotypic covariation.
- *Epigenetic PCA analysis.* To understand the genetic and environmental factors influencing genome regulation in our population, we also examined the relationship of 186 exposures (**Supplementary Table 6**) with the principal components of variation in methylation at the 16,444 CpG sites that are differentiated between our Asian ethnic groups. We used PCA as a data reduction strategy to identify the primary axis of variation in the methylation at these CpG sites. We then tested the associations with the 5 PCs with potential exposure, adjusted for age, sex, ethnicity, methylation array control probe PCs, and white cell subset composition estimated by the Houseman method^29^. We again inferred statistical significance at P<0.05/32, based on an estimate of 32 independent phenotypes derived from PCA of phenotypic covariation.
- *Metabolic variation*. To explore the variation in metabolite levels across ethnicities, we randomly split the dataset into discovery (70%) and test (30%) cohorts. Using linear regression analysis, we estimated the association between variation in levels of 1,073 metabolites and self-reported ethnicities (Malay compared to Chinese, Indian compared to Chinese, and Malay compared to Indian) in the discovery cohort, adjusted for age, sex, and shipment batch. We applied a Bonferroni-corrected p-value threshold of 1×10^-5^ to account for multiple testing (1,073 metabolites x 3 pair-wise tests). We then repeated the same set of analyses for these 162 metabolites in the replication cohort, and a subset of 153 metabolites that met the following criteria: 1) significantly associated with ethnicity in the discovery cohort at P<1×10^-5^, and 2) significantly associated with ethnicity in the test cohort at P<0.05 and with the same direction of estimates. In an age and sex-matched cohort of 1,146 participants per ethnicity, we performed PCA of the 153 metabolites to assess the extent of clustering of individuals by ethnicity. Out of these 153 metabolites, 128 were well-characterized and known metabolites. We evaluated associations between these 128 metabolites and four common health outcomes: hypertension, obesity, T2D, and CVD, using logistic regressions adjusted for age, sex, and shipment batch. For each phenotype, we applied a Bonferroni-corrected p-value threshold of 4.7×10^-5^ to account for multiple testing (1,073 metabolites). We also evaluated associations between a metabolite of interest (1-margaroyl-2-arachidonoyl-GPC) and FFQ foods, adjusted for age, sex, ethnicity and shipment batch, and reported Bonferroni-Hochberg corrected p-values for the top four foods. Furthermore, for each of these metabolites, we calculated partial R-squared values to estimate the contribution of genetic ancestry and various demographic and lifestyle factors on metabolic variation. Genetic ancestry was represented using the first 50 genetic PCs, and dietary habits using the first 10 PCs representing 169 food items and major macronutrients. Finally, pairwise correlation between metabolites was estimated using Pearson correlation and a significance p-value threshold of 1×10^-6^ was applied to account for multiple testing. Metabolites were grouped into 10 categories (super-pathways) and annotated to pathways or chemical classes based within each category (sub-pathways). The circos plot was generated using the *circlize* package.
- *RNA sequencing.* Expression quantitative trait loci (eQTLs) were analysed using Matrix eQTL (R package *MatrixEQTL*), with gene expression modelled as a regression model of genotypes and covariates, including age, sex, ethnicity, RIN (RNA integrity number) and the top 6 PEER (Probabilistic Estimation of Expression Residuals) factors.^54^ For the identification of significant *cis* and *trans* eQTLs, a Bonferroni-corrected p-value threshold at P<0.05 was applied.
- *Genome Wide Association Studies (GWAS)*. To identify genetic variants associated with metabolite levels in the HELIOS dataset, we first divide the cohort to select only individuals having metabolite data and were clustered in our three main ancestry groups (Chinese, N=5,961; Indian: N=1,470; Malay: N=838) that were determined by our data driven approach. The individuals in the three admixed group (n = 409) were not included in the analysis. We then perform GWAS QC and analysis for each group separately, followed by inverse variance meta-analysis to create summary statistics across the study population. GWAS variant QC filters were: MAF < 0.5%, HWE p-value <1×10^-6^, Missingness <2%. Sample filters were pi_hat < 0.75, IBC <|0.2| and Sex-mismatch. We used PLINK2^27^ to get the final set of samples and variants to be used for the analysis. Overall, 5,940 Chinese, 1,461 Indians and 833 Malays with 12.7, 16, and 14.7 million variants respectively were included in the analysis. For the GWAs of metabolites, we log transformed the metabolite data and removed individuals with the highest deviation (>5 SD from the mean). Age, sex, top 20 genetic PCs, and batch were used as covariates in the analysis. The individual GWAS for each ancestry was performed using REGENIE.^55^ The subset of SNPs for REGENIE step 1 were chosen after filtering for MAF <5%, HWE P<1×10^-6^ in the ethnic group being analysed. We removed the MHC and chr8 inversion regions, followed by LD pruning at an r^2^ of 0.05 within a 200kb window. Meta-analysis of the three summary statistics was performed using METAL with a fixed effect model controlling for genomic inflation across each dataset. Variants were filtered for being in at least two datasets, heterozygosity P>0.05 and max difference between allele frequencies <0.5.
- *SMR and colocalization:* Summary data-based mendelian randomization analysis (SMR)^56^ was performed to identify pleiotropic association between gene expression (exposure) (from the eqtlgen dataset^56^) and metabolite levels (outcome) using GWAS summary statistics. To limit the number of tests, we include SNPs that pass genome-wide significance in our GWAS as well as in the cis-eQTL dataset. Analysis was performed using the SMR tool^57^. For the metabolite-gene pairs with significant SMR association, we performed colocalization analysis using the *coloc* package implemented in R.^58^ The region of 1MB on each side of the SMR associated SNP was used for colocalization analysis under a single causal variant assumptions and the default prior probabilities. Metabolite-Gene pairs with a coloc H4 posterior probability >0.7 were considered to be colocalized and share a common causal variant.
- *Validity and reproducibility assessment of measurements*. Pairwise correlation matrix across phenotypes was calculated using Pearson correlation analyses for z-scored transformed measurements. The reproducibility of 107 measurements in 10 domains (**Supplementary Table 14**) between baseline test and the repeated study was assessed using correlation coefficients calculated from Spearman correlation analysis for z-scored transformed measurements.

## Data availability

The HELIOS phenotype and genotype data used in this manuscript are protected and are not publicly available due to data privacy regulations. Data access request can be submitted to the HELIOS Data Access Committee by emailing for details. For accessing de-identified National Health and Administrative records linked through TRUST, please contact TRUST platform (https://trustplatform.sg) for details.

## Code availability

The analytic codes are available in the following github repository.

## Acknowledgements

This study is supported by Singapore Ministry of Health’s (MOH) National Medical Research Council (NMRC) under its OF-LCG funding scheme (MOH-000271-00), Singapore Translational Research (StaR) funding scheme (NMRC/StaR/0028/2017), the National Research Foundation, Singapore through the Singapore MOH NMRC and the Precision Health Research, Singapore (PRECISE) under the National Precision Medicine programme (NMRC/PRECISE/2020) and intramural funding from Nanyang Technological University, Lee Kong Chian School of Medicine and the National Healthcare Group. RNA sequencing was partially funded by i) Ministry of Education Academic Research Fund Tier 1 Grant (RS09/20), ii) A*STAR-NHMRC Joint Grant Call (A20PRb0138), iii) Start-Up Grant (awarded to M.Loh [PI]) from Lee Kong Chian School of Medicine, Nanyang Technological University, Singapore and iv) Imperial - Nanyang Technological University Collaboration Fund (awarded to M.Loh [PI]). T.M. was funded by Dean’s Postdoctoral Fellowship from the Lee Kong Chian School of Medicine.

This study made use of data generated by Ministry of Health (MOH) and Immigration and Checkpoints Authority (ICA). This study was supported by the Trusted Research and Real-World-Data Utilisation and Sharing Tech platform (“TRUST Platform”) developed by the Ministry of Health and Smart Nation and Digital Government Office, through the use of its research data analytics facilities. The views expressed are those of the author(s) are not necessarily those of the Government, MOH and ICA investigators or institutional partners. The computational work for this study was partially performed on resources of the National Supercomputing Centre, Singapore (https://www.nscc.sg).

We thank all participants and research staff who made the study possible. We thank Tomas Gonzalez, Lewis Griffiths, Stefanie Hollidge, and Antony Siahaan (MRC Epidemiology Unit, University of Cambridge), Ian Goon and Samiul Hoque for assistance in accelerometry data extraction and analysis, Mr. Shaikh Fairul Edros Shaikh Ahmad (Earth Observatory Singapore, NTU) for guidance in geospatial data analysis, and Clare Whitton for the collaborative development of e-FFQ and the data collection platform.

## Author contributions

J.C.C. P.E, E.R, J.N, L.E.S, J.L, T.M., K.T, and L.L, L.T.H, and J.B., conceived and designed the HELIOS study. T.M., T.T.Y.Y., C.W.L., K.S.K., L.G.L., B.L.C.C., R.D., G.W. and Y.Y.W implemented the study and collected data. T.M., N.S, D.L.Y.W., P.R.J, D.T., G.A.M, K.E.W, P.A.S, L.P.Y., Y.Z.X., N.B., C.B., M.H., P.G., E.J.L, S.B and H.S. curated epidemiological and molecular data. X.W., T.M., N.S., P.R.J., H.K.N., D.L.Y.W., D.T., R.D., M.Lam, and M.Loh performed the data analyses. J.C.C. supervised the study implementation, data curation and analyses. X.W., T.M., N.S., P.R.J., H.K.N., D.L.Y.W., M.Lam, M.Loh, and J.C.C. wrote the manuscript. All authors reviewed and contributed to the revision of the submitted manuscript.

## Additional Information

Supplementary Information is available for this paper.

Correspondence and requests for materials should be addressed to John C. Chambers.

Reprints and permissions information is available at www.nature.com/reprints.

## Supplementary

**Extended Table 1.** Demographics, lifestyle, physiological, and molecular measurements of participants at baseline, HELIOS 2018-2022.

| Characteristics | Chinese<br>N = 6,784 | Indians<br>N = 1,807 | Malays<br>N = 1,354 | Overall <sup>a</sup><br>N = 10,004 |
| --- | --- | --- | --- | --- |
| Age at baseline (years), mean (SD) | 53.2 (12.0) | 53.1 (11.5) | 51.0 (11.1) | 52.9 (11.8) |
| Male, % | 2658 (39.2) | 858 (47.5) | 483 (35.7) | 4027 (40.3) |
| Marital status, % |  |  |  |  |
| Single | 1433 (21.3) | 187 (10.4) | 143 (10.7) | 1772 (17.9) |
| Married | 4717 (70.1) | 1371 (76.5) | 1026 (76.6) | 7159 (72.2) |
| Separated, divorced, widowed | 577 (8.58) | 234 (13.1) | 171 (12.8) | 986 (9.9) |
| Education, % |  |  |  |  |
| Secondary school or less | 1468 (21.7) | 562 (31.2) | 563 (41.7) | 2606 (26.1) |
| Post-secondary <sup>b</sup> | 1920 (28.4) | 484 (26.9) | 509 (37.7) | 2925 (29.3) |
| Bachelor | 2420 (35.8) | 457 (25.4) | 208 (15.4) | 3100 (31.1) |
| Postgraduate | 957 (14.1) | 299 (16.6) | 69 (5.11) | 1344 (13.5) |
| Homeowner, % | 5878 (87.2) | 1550 (86.1) | 1138 (84.4) | 8614 (86.6) |
| House type, % |  |  |  |  |
| Public housing | 4392 (65.0) | 1384 (76.9) | 1248 (92.5) | 7056 (70.8) |
| Private housing | 2340 (34.6) | 407 (22.6) | 94 (6.97) | 2867 (28.8) |
| Other | 27 (0.4) | 9 (0.5) | 7 (0.5) | 44 (0.4) |
| Household monthly income ≥ \$10,000 <sup>c</sup> | 1843 (32.0) | 325 (20.4) | 129 (11.0) | 2318 (27.0) |
| Cigarette smoking, % |  |  |  |  |
| Never | 5078 (75.3) | 1260 (70.0) | 818 (60.8) | 7188 (72.2) |
| Past | 1261 (18.7) | 347 (19.3) | 288 (21.4) | 1917 (19.3) |
| Current | 405 (6.0) | 194 (10.8) | 240 (17.8) | 845 (8.5) |
| Alcohol consumption, % |  |  |  |  |
| Never | 5260 (78.1) | 1364 (75.8) | 1311 (97.5) | 7968 (80.2) |
| 1-3 times per month | 695 (10.3) | 173 (9.6) | 13 (1.0) | 890 (9.0) |
| 1-4 times per week | 668 (9.9) | 229 (12.7) | 17 (1.3) | 929 (9.4) |
| Daily | 108 (1.6) | 34 (1.9) | 3 (0.2) | 146 (1.5) |
| Physical activity (MET-h/day), median (IQR) | 7.23 (22.8) | 17.3 (30.6) | 19.6 (30.5) | 9.68 (26.0) |
| Sedentary time (h/day) <sup>d</sup>, mean (SD) | 6.04 (2.98) | 5.54 (2.97) | 5.51 (3.11) | 5.86 (3.01) |
| BMI (kg/m<sup>2</sup>), mean (SD) | 23.6 (3.81) | 27.1 (4.93) | 28.2 (5.50) | 24.9 (4.68) |
| < 18.5 | 341 (5.0) | 24 (1.3) | 14 (1.0) | 380 (3.8) |
| 18.5 to < 23.0 | 2962 (43.8) | 323 (17.9) | 184 (13.6) | 3486 (34.9) |
| 23.0 to < 27.5 | 2494 (36.9) | 717 (39.7) | 488 (36.1) | 3722 (37.3) |
| ≥ 27.5 | 965 (14.3) | 740 (41.0) | 667 (49.3) | 2390 (24.0) |
| Waist circumference, mean (SD) | 80.6 (10.6) | 89.8 (11.9) | 88.1 (12.4) | 83.3 (11.8) |
| Total fat percentage <sup>e</sup>, mean (SD) | 36.6 (6.70) | 41.0 (7.61) | 41.3 (7.18) | 38.0 (7.27) |
| VAT volume (cm<sup>3</sup>) <sup>e</sup>, mean (SD) | 629 (275) | 826 (296) | 791 (309) | 687 (296) |
| Handgrip strength (kg), mean (SD) |  |  |  |  |
| Left | 26.2 (8.58) | 26.8 (8.76) | 25.2 (8.71) | 26.2 (8.65) |
| Right | 28.7 (9.29) | 29.1 (9.48) | 27.5 (9.31) | 28.6 (9.36) |
| Blood pressure (mmHg), mean (SD) |  |  |  |  |
| Systolic | 121 (18.0) | 123 (18.6) | 123 (18.4) | 121 (18.2) |
| Diastolic | 75.4 (10.4) | 77.0 (10.6) | 75.9 (10.4) | 75.7 (10.5) |
| Molecular, mean (SD) |  |  |  |  |
| Triglycerides (mmol/l) | 1.21 (0.76) | 1.39 (0.89) | 1.43 (0.97) | 1.27 (0.82) |
| Total cholesterol (mmol/L) | 5.26 (0.96) | 5.01 (0.98) | 5.31 (1.07) | 5.22 (0.99) |
| HDL (mmol/l) | 1.61 (0.42) | 1.30 (0.33) | 1.42 (0.37) | 1.53 (0.42) |
| LDL (mmol/l) | 3.10 (0.84) | 3.09 (0.86) | 3.24 (0.95) | 3.12 (0.86) |
| HbA1c (%) | 5.60 (0.65) | 6.06 (1.23) | 5.95 (1.21) | 5.73 (0.89) |
| Fasting plasma glucose (mmol/l) | 4.95 (0.91) | 5.51 (1.79) | 5.35 (1.86) | 5.11 (1.29) |
| Insulin (U/L) | 9.25 (6.46) | 14.4 (10.8) | 12.0 (8.27) | 10.6 (7.94) |
| CRP (mg/dL) | 1.29 (3.94) | 3.41 (5.60) | 2.93 (6.30) | 1.90 (4.73) |

Abbreviations: CRP: C-Reactive Protein; DEXA: dual energy X-Ray absorptiometry; HbA1c: haemoglobin A1c; HDL: high-density lipoprotein; IQR: interquartile range; LDL: low-density lipoprotein; MET: metabolic equivalent task; SD: standard deviation; T2D: type-2 diabetes.
<sup>a</sup> Included additional 59 participants with other ethnicities.
<sup>b</sup> Included junior college or IB or equivalent, vocation or the Institute of Technical Education, or diploma.
<sup>c</sup> Included salary, rental income, investments, pensions, and government transfers.
<sup>d</sup> Calculated as average sitting hours per day including weekends.
<sup>e</sup> Derived from DEXA whole-body scan.
Notes: All calculations were based on participants with available information. Participants with missing values on any one characteristic were not counted in the calculation of the respective characteristic. The percentages might not add to 100 due to rounding.

**Extended Table 2.** Spectrum of baseline measurements and biological samples in the HELIOS.

| Domain | Components |
| --- | --- |
| Health and lifestyle questionnaire | Demographics and socioeconomic status; tobacco smoking; alcohol consumption; mobile-phone use; Pittsburgh Sleep Quality Index; Patient Health Questionnaire-9; Generalized Anxiety Disorder-7; childhood and early life history; personal health history; reproductive health history; International Physical Activity Questionnaire;<sup>26</sup> nurse questionnaire (medication and hospitalization history) |
| Dietary intake | Electronic Food Frequency Questionnaire on a range of food and beverages frequently consumed in multi-ethnic Singapore<sup>31</sup> |
| Cognitive health | UK-Biobank based computerized cognitive test comprising reaction time, numerical recall, fluid intelligence (verbal and numerical reasoning), paired associate learning, episodic memory, and Stroop test<sup>32</sup> |
| Physiological measurements | Anthropometry (height, weight, body impedance); waist and hip circumference; blood pressure; ECG; hand grip strength; arterial stiffness assessments; dermatological assessments; audiological assessments; exercise treadmill test |
| Imaging | 3D carotid ultrasound; OCT/ OCTA retinal imaging; colon-fundus photography; DEXA scan (hip, lumbar and whole-body) |
| Routine biochemistry (fasted) | Lipid panel; glucose; HbA1c; insulin; renal panel; blood panel; C-Reactive protein; Vitamin D; Follicle-Stimulating Hormone; Luteinizing Hormone; Oestrogen |
| Biological samples | Blood in EDTA, LH, clot activator, and acid citrate dextrose vacutainers; urine; saliva; stool (subset); skin tape |

Abbreviations: DEXA: dual energy X-Ray absorptiometry; ECG: electrocardiogram; EDTA: ethylenediamine tetra-acetic acid; LH: lithium heparin; OCT: optical coherence tomography; OCTA: optical coherence tomography angiography.

**Extended Figure 1.**
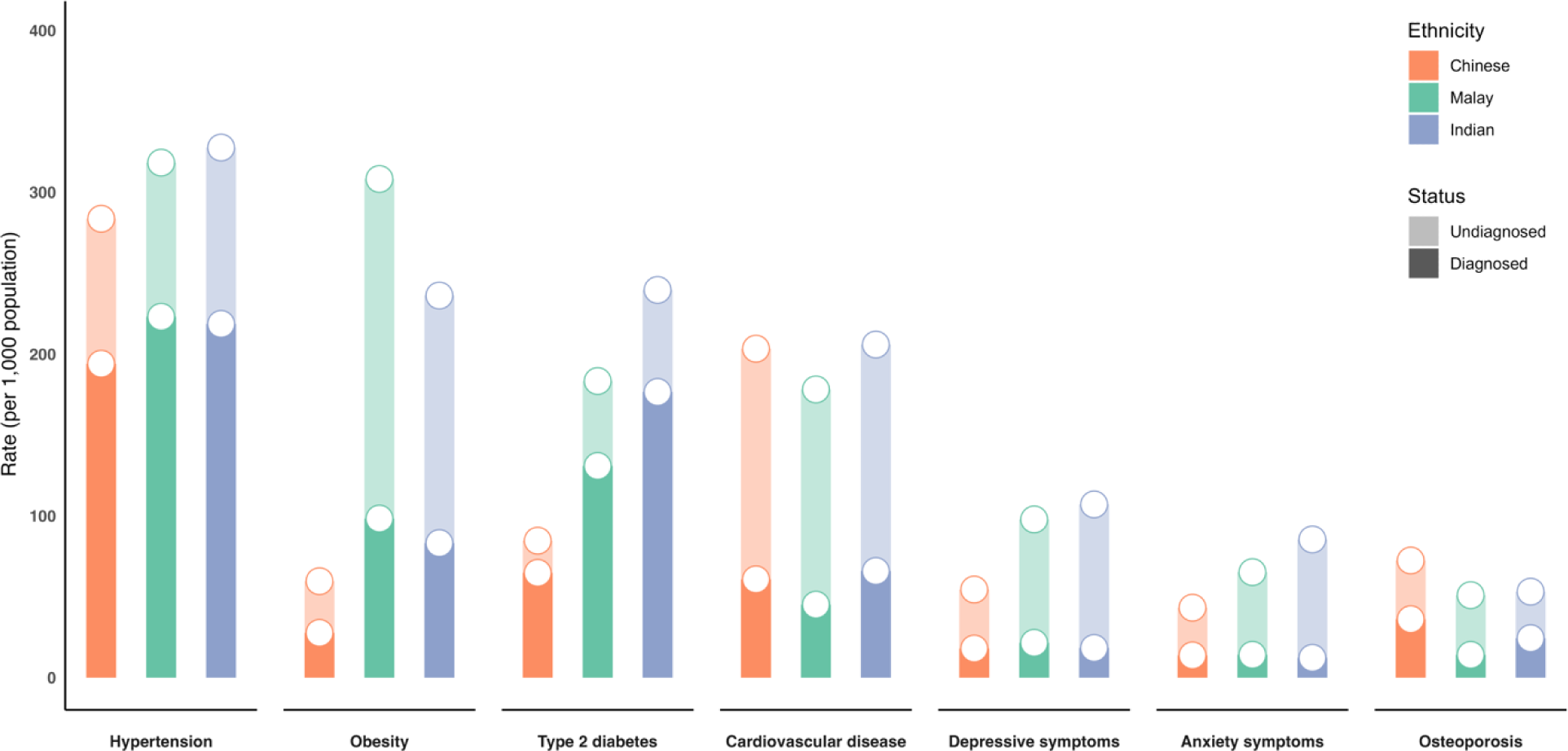
Divergent health states across three Asian population groups in Singapore. Diagnosed vs. undiagnosed cases were defined as the participants with self-reported records of diagnoses or the use of pharmacological treatment. Undiagnosed cases were defined as those who did not self-report the conditions but met the following criteria: Hypertension: blood pressure ≥140/90 mmHg; Obesity: BMI ≥30 kg/m^2^; Type 2 diabetes: fasting glucose ≥7.0mmol/L or HbA1c ≥6.5%; Cardiovascular disease: subclinical atherosclerosis defined as presence of atherosclerotic plaque or mean cIMT ≥0.8; depressive symptoms: PHQ-9 score ≥10; Anxiety symptoms: GAD-7 score ≥10; Osteoporosis: lumbar spine bone mineral density T-score of -2.5 or below. Abbreviations: BMI: body mass index; cIMT: carotid intima-media thickness; GAD-7: Generalised Anxiety Disorder Assessment-7; PHQ-9: Patient Health Questionnaire-9.

**Extended Figure 2.**
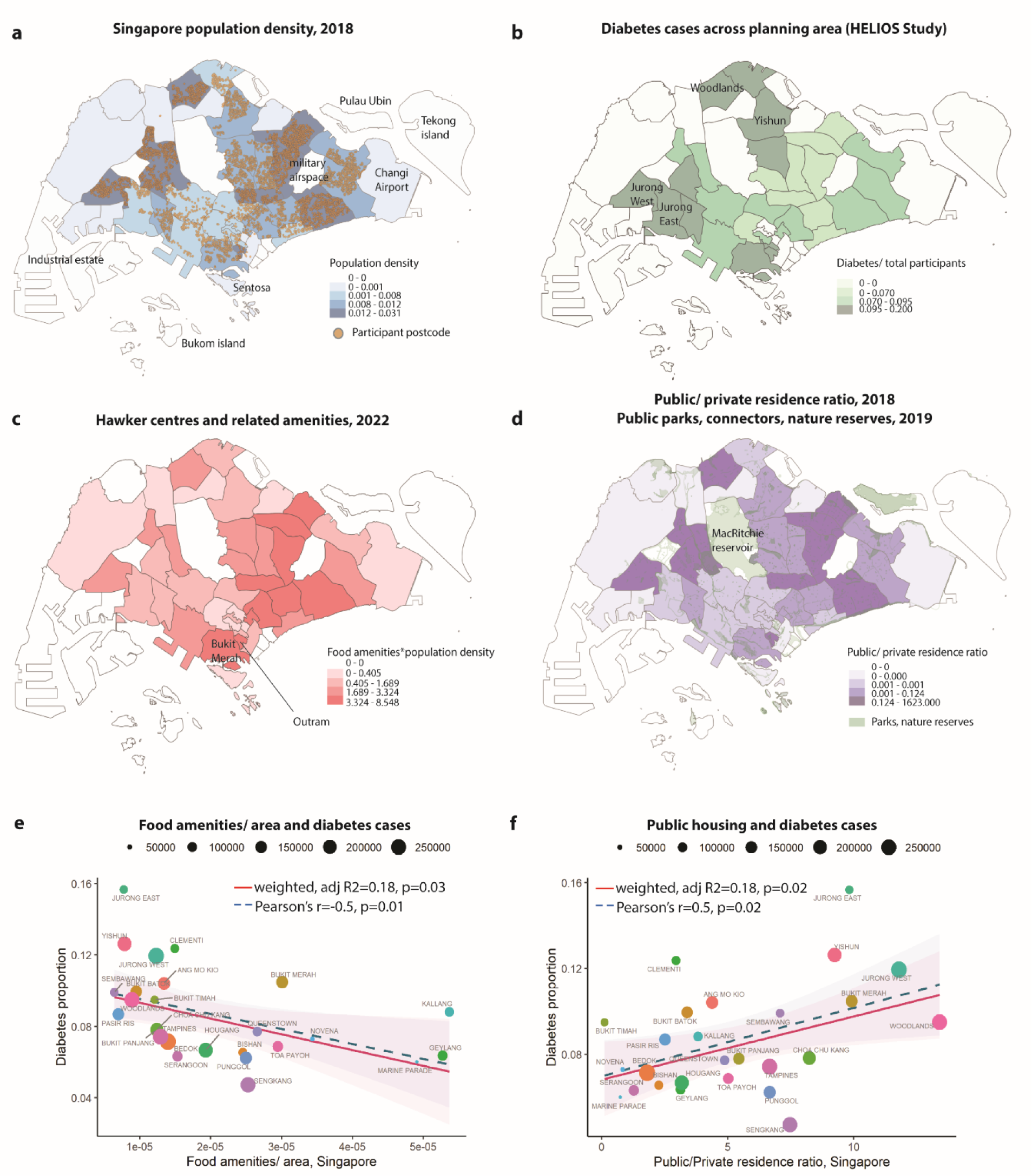
Environment exposure across three populations. **a)** The Singapore population density across planning area overlayed with HELIOS study participant postcode; **b)** The distribution of diabetes cases; **c)** The distribution of food amenities, including hawker centres, shopping malls, restaurants, and convenience stores; and **d)** the ratio of public/ private residence, overlayed with public parks, connectors, and nature reserves. The built environment has impact health outcomes, as illustrated by the correlations of diabetes case proportions and **e)** the density of food amenities/ area, and **f)** the public/private residence ratio, weighted by population numbers.

**Extended Figure 3.**
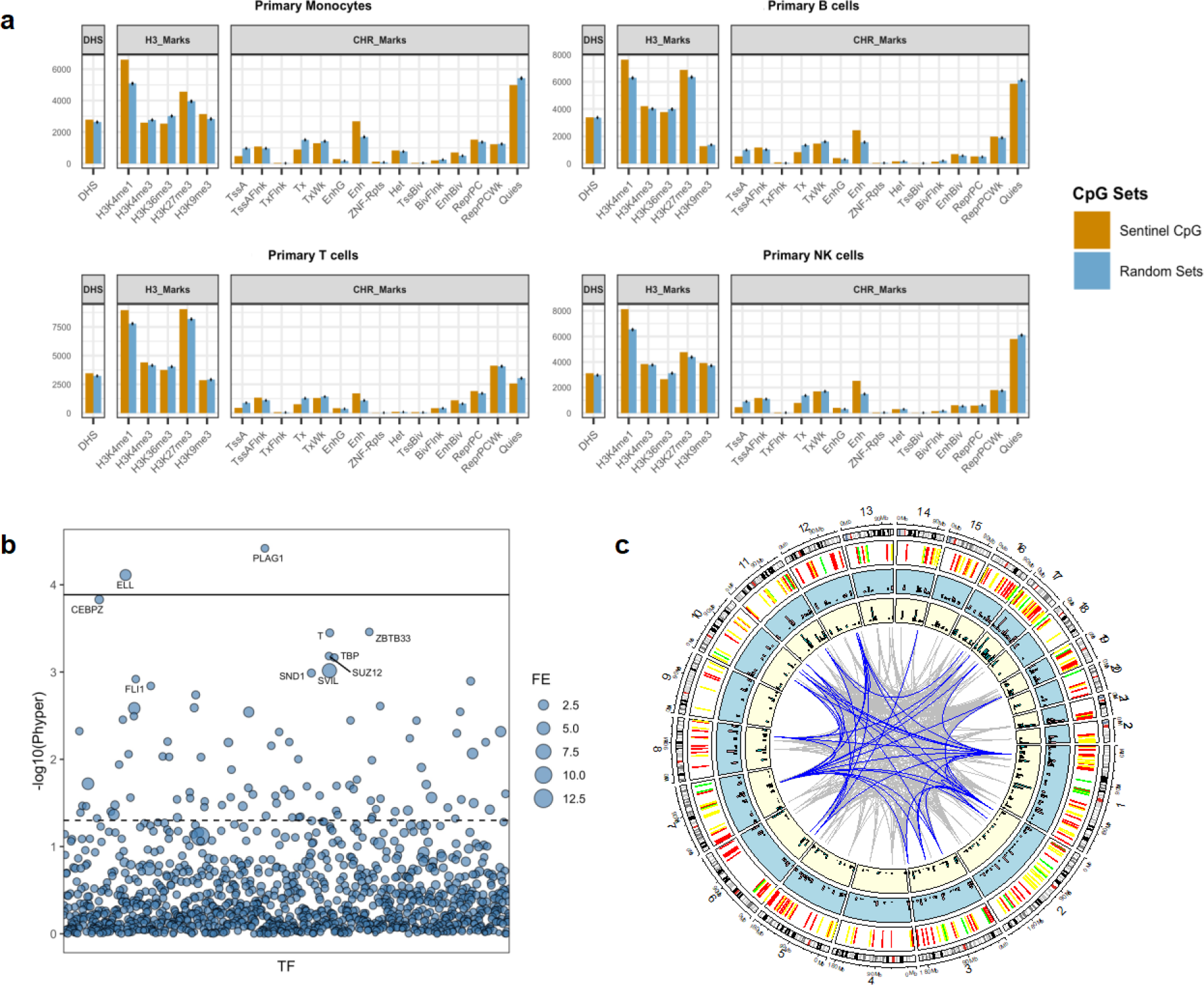
Epigenetic variation between Asian populations. **a)** Functional annotation and enrichment of ethnically differentiated CpGs across 4 blood cell types. Enrichment is shown as observed count vs expected background count across DNase 1 Hotspots (DHS); five Histone 3 marks and 15 Chromatin States. **b)** Enrichment of ethnically differentiated CpGs across 1210 transcription factors (TFs) from the ReMAP database. The size of the circle represents the fold enrichment compared to background. **c)** Circos plot of the epigenome-wide association of DNA methylation in blood with incident T2D and BMI, along with methylation profile by ethnic populations and pairwise correlation between CpGs. Chromosome numbers and base positions are shown on the outermost ring. The second ring illustrates the ethnic group with the most unfavourable average methylation levels (i.e., corresponding to highest risk for T2D; Chinese: Green, Malay: Yellow; Indian: Red) of 314 sentinel CpG sites that predict T2D [P<8.62×10^-8^]. The next two rings show the CpG-specific association test results [-log10(P); axis starts at P=1×10^-22^] ordered by genomic position (light yellow: incident T2D in TOAST; light blue: BMI in HELIOS). The innermost connections summarise the pairwise correlations between sentinel CpG sites (Gray: |Correlation| > 0.5, Blue: |Correlation| > 0.6).

**Extended Figure 4.**
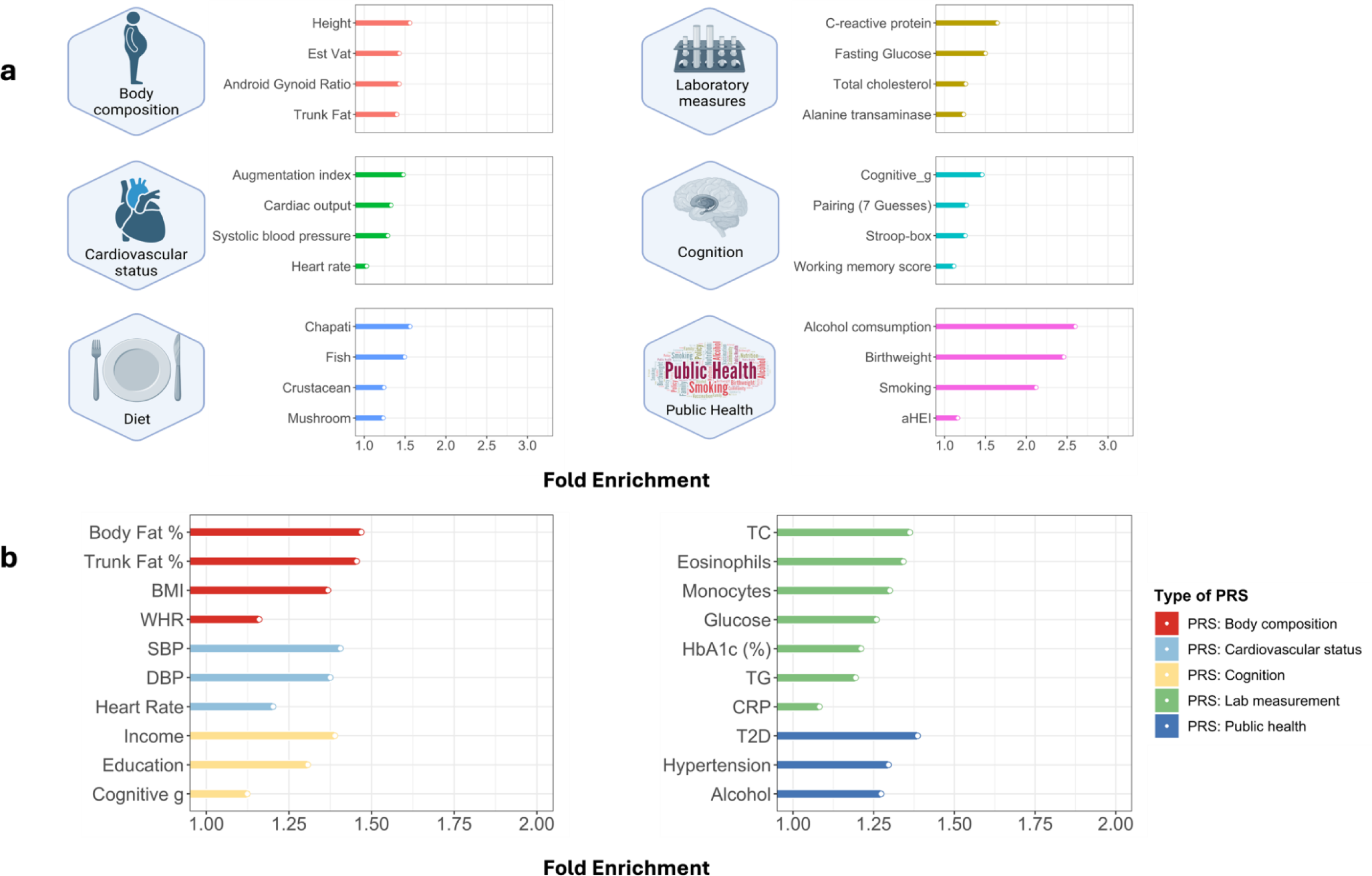
Enrichment of ethnically differentiated CpGs across clinical, behavioural, and genetically inferred traits. Enrichment of ethnically differentiated CpGs for association with directly measured clinical traits (Panel a) and genetically inferred exposures based on PRS (Panel b). All enrichment tests are significant after multiple testing correction.

**Extended Figure 5.**
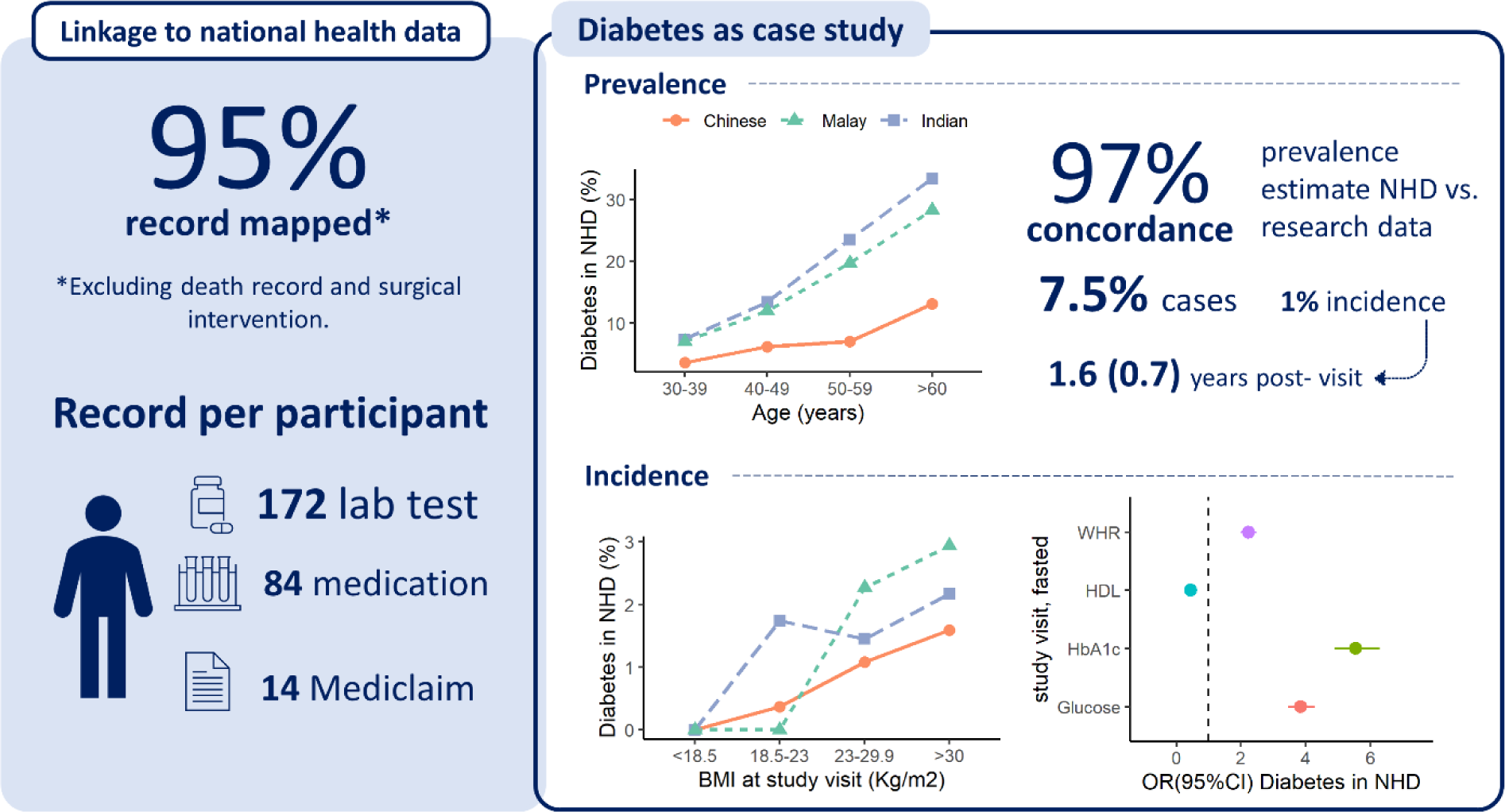
Infographics of national health data linkage. NHD: national health data. HDL: High-density lipoproteins. WHR: Waist-hip ratio.

**Extended Figure 6.**
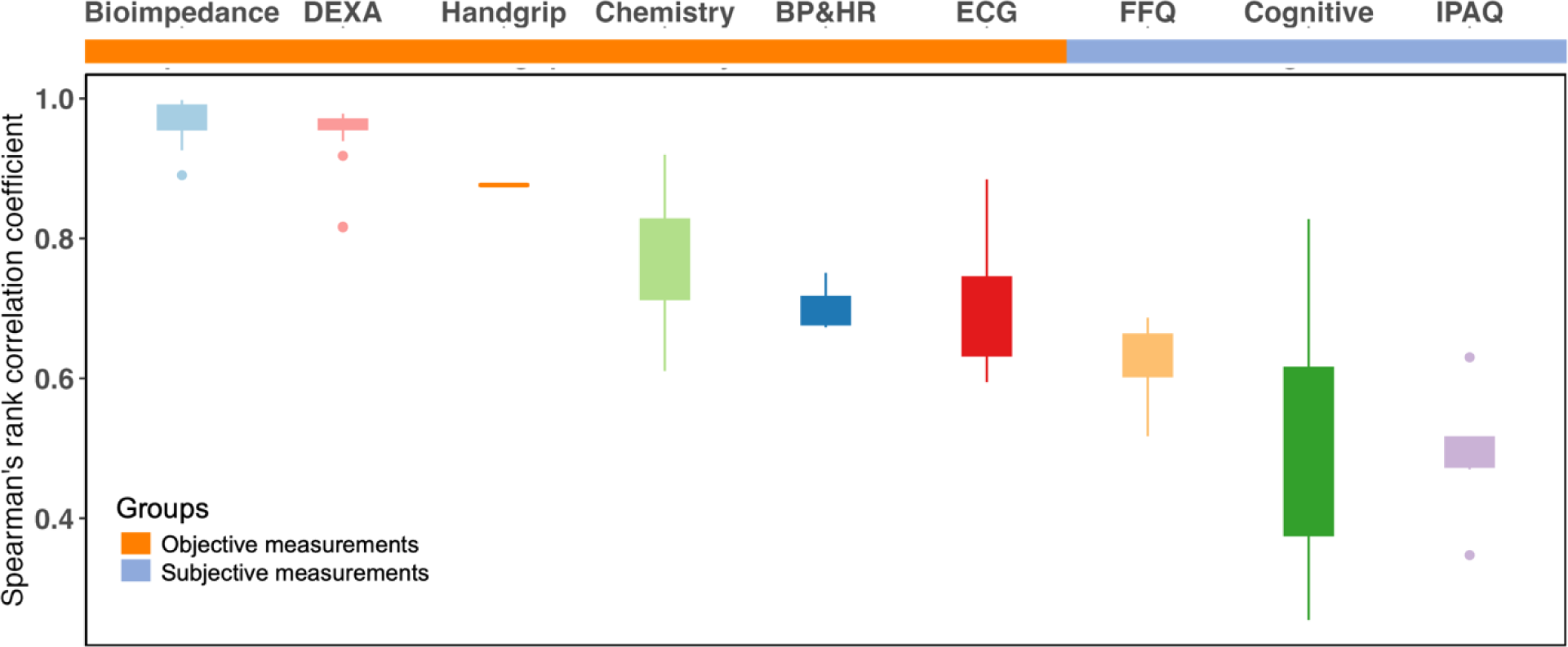
Reproducibility of measurements in HELIOS. Spearman correlation analyses were used to calculate the correlation coefficients between z-scored baseline and retest measurements (N= 398). The measurement components for each group and the total number of variables per group were detailed in **Supplementary Table 4**. Abbreviations: BP: blood pressure; DEXA: dual-energy X-ray absorptiometry; ECG: electrocardiogram; FFQ, food frequency questionnaire; HR: heart rate; IPAQ: International Physical Activity Questionnaire.

## Supplementary Methods

### Questionnaire

A combination of self-administered and nurse-administered questionnaires were applied to collect information on demographics, socioeconomic status, physical activity, tobacco smoking, alcohol consumption, diet, mobile phone use, sleep, mood, childhood and early life history, personal health status and disease history, reproductive history, cognitive function, medications, and health supplements. Physical activity was assessed using the International Physical Activity Questionnaire long form, which documented the type, frequency, and duration of various activities in the domains of transportation, occupation, leisure time and household in the last 7 days. Dietary intake (servings/day) was assessed using electronic Food Frequency Questionnaire (FFQ) on a range of food and beverages frequently consumed in multi-ethnic Singapore. Both questionnaires were validated in adult Singapore population.^42,43^ Sleep was assessed using the Pittsburgh Sleep Quality Index^59^, and mood was evaluated with Patient Health Questionnaire (PHQ-9)^38^ and Generalised Anxiety Disorder (GAD-7)^39^. Cognitive function comprising six computerized components was performed based on UK Biobank cognitive test, which was proved with substantial validity and reliability for some test components.^60^

### Physiological measurements

Body weight and height were measured once using computerized measuring instruments with automated data capture. Chest, waist, hip circumference, and leg length were measured using a non-stretchable sprung measuring tape and manually entered in the IT system which could automatically highlight impossible or implausible values. Body fat composition by bioimpedance were measured using an Inbody 770 device or equivalent. Systolic and diastolic blood pressures were measured 3 times in the right arm using an Omron HEM-9210T (or equivalent) blood pressure monitors. Right- and left-hand grip strengths were measured using a Jamar Plus Hand Dynamometer (or equivalent). Skin physiology measurement includes trans-epidermal water loss measured by a vapometer, surface hydration measured by a MoistureMeterSC and skin surface pH measured by a pH meter. Lung function was assessed using spirometry via MIR Spirolab monitor (or equivalent). Participants were asked to provide up to 4 recordings to overcome the learning effect. Cardiac evaluation by 12-lead electrocardiogram (ECG) recorded using a GE Healthcare CASE device (or equivalent) according to published international standards^61^ was performed to identify a wide variety of cardiac abnormalities, such as “silent” myocardial infarction, arrhythmia, and left ventricular hypertrophy. Arterial stiffness was measured using the VICORDER, a non-invasive device that allows pulse wave velocity to be measured simply and rapidly, or equivalent. 3-D carotid ultrasound scans were performed on both left and right carotid arteries with participant recumbent at 45 degrees using a Philips EPIQ 7 (or equivalent) device to assess carotid plaque area and volume and identify subclinical atherosclerosis. Ophthalmology assessment was performed using optical coherence tomography angiography (OCTA), optical coherence tomography (OCT), and colour fundus photography. Visual acuity, refraction, intra-ocular pressure, and corneal biochemical properties were also assessed. All measurements will be made with the participant in a sitting position in a darkened room, according to internationally recognised protocols. Physical fitness was evaluated by a sub-maximal treadmill test comprised of a walk at speed of 4km/h, 5km/h and 6 km/h for 2 minutes each, and a 3% followed by a 6% increase in gradient at 6km/h for 2 minutes each. Heart rate was monitored throughout the test. Blood pressure was measured at the beginning and end of the treadmill test. A continuous 3-lead ECG was captured for non-diagnostic purposes to provide additional cardiovascular phenotypes. Wrist-based accelerometer devices were worn for 7-days to monitor physical activity. Dual energy X-ray absorptiometry (DEXA) scan for whole body, hip and lumbar spine was used to measure bone mineral density and body composition. Audiological assessments were performed via tympanometry twice per ear, followed by pure-tone audiometry once.

### Collection and storage of biological materials

Blood was collected from all participants at a single time point by a certified phlebotomist during the baseline assessment visit and processed according to well established protocols, complying with international best practice. The blood samples were either analysed immediately for haematology, coagulation and biochemistry tests or stored at -80°C for future research use. A morning sample of middle stream urine and 24h fasting saliva were also be collected for each participant. Stool collection is optional. The stool samples were collected in a specimen container with lid tightly closed. The urine and saliva samples were immediately placed on ice and aliquoted within half an hour of collection and stored in -80°C freezers. Two skin tapes per body site were collected using aseptic technique by tape stripping the antecubital fossa, upper back, and volar forearm. These tape specimens do not require immediate processing but will be stored at -80°C as soon as possible but no longer than 30 minutes. All biological samples were processed manually, by one or two technicians using a Laboratory Information Management System. Biological samples were prepared and stored in the freezers of Nanyang Technological University Lee Kong Chian School of Medicine.

### Quality management

To guarantee a high-quality data, a proven and comprehensive study IT system developed by Imperial College London with computerized data entry, direct equipment interface and automated alarming system for impossible or implausible values were employed to prevent manual data input errors. The computerised direct data entry facilitates the collection of accurate and complete data by allowing internal quality check, automated coding, and immediate access. Where participants are unable to complete the computerised questionnaire, an adult relatives or trained staff assisted in the completion (no assistance was allowed in the cognitive test). This system has successfully supported the Qatar Biobank study for delivering an 18-month pilot study with over 2,500 participants. For physiological measurements, all examinations were undertaken by trained and accredited nurses or technicians under internationally accepted protocols using standardised instruments and overseen by clinical scientists. The data management teams will also assess the completeness and quality of the data on a regular basis. A quality control report will be generated and discussed internally.

### Return of findings

The HELIOS is committed to return assessment results and written feedback to all participants. The baseline screening data in the format of structured report, including height, weight, waist circumference and body composition e.g. body fat percentage, blood pressure, ECG, DEXA scan, full blood count, lipid profile, glucose and HbA1c, uric acid, active smoking status, self-reported medical history and list of medications, along with a booklet explaining the meaning of the tests and the interpretation of the results within 4 weeks of the assessment date. In addition, a written protocol is established to describe what represents a clinically significant abnormality requiring clinical action. Participants with non-urgent clinical findings will receive a report within approximately four weeks to advise them to see their own doctor for further advice. Major clinical findings are discussed with a senior NHG physician on the same day for a decision on appropriate action. Clinical findings of immediate significance will be referred immediately to Accident & Emergency (A&E). All other measurements conducted in the course of the HELIOS study are being done for research purposes only and will not therefore be fed back to participants routinely.

### Ethics and data security

The HELIOS study fully complies with the Personal Data Protection Act (PDPA) requirements.^62^ Use of the research data and samples is regulated by the HELIOS Study “Scientific and Data Access Committee”. Permission to Access to use the data is evaluated based on written applications, according to scientific merit. All research data are de-identified. The personal details of participants are stored separately from the research database to enhance data security. The codes that match the personal and research identifiers are held separately from both the personal and research data.

